# Machine Learning-Based Prediction of Maternal Morbidity across Heterogeneous Populations in the United States using Sequential Modeling of the *All of Us* Dataset

**DOI:** 10.64898/2026.08.25.26360552

**Authors:** Haojun Zhuang, Arthurine Zakama, Katherine Heller, Shakeela Faulkner, Bruce Gollub, Nichole Young-Lin, Irene Y. Chen, Mercy Asiedu

## Abstract

In this work, we demonstrate the unprecedented value of NIH’s “All of Us Research Program” (AoURP) dataset in studying maternal morbidity and building predictive machine learning (ML) models across heterogeneous populations in the United States. We developed robust and data-driven preprocessing pipelines to curate a longitudinal, multi-site, multimodal, and demographically diverse pregnancy dataset (20,253 subjects; 27,525 pregnancy episodes) from AoURP data, using electronic health records (EHR) (Conditions, Labs, Measurements) and survey responses (Social Determinant of Health (SDoH)), focusing on 7 crucial maternal health adverse outcomes. After characterizing data quality, missingness, and heterogeneity, we performed statistical correlation analysis to identify risk factors. We subsequently developed XGBoost and sequential LSTM models to predict the adverse outcomes, reaching state-of-the-art performance for multiple outcomes. We conducted model interpretability post-hoc analysis to understand success points and fairness analysis to evaluate implications for socio-economic disparities. Four practicing physicians reviewed the set of statistically significant and ML model identified features to assess their clinical validity and novelty. Most features identified through either statistical correlations or ML feature importance analysis aligned with known clinical risk factors. Several features were identified that the ML models used but that are not currently used in clinical practice and may merit further clinical investigation. Fairness analysis revealed certain associations with SDoH and age highlight areas that warrant continued monitoring. Overall, we demonstrate that meaningful populational level patterns can be extracted, and high-performing machine learning models can be trained on this longitudinal, diverse, multi-site dataset. Important risk features, particularly novel ones identified, if validated, could inform new strategies for maternal care or enable development and validation of outcome-specific, clinically deployable ML models.

## Introduction

Maternal health in the United States represents a critical public health crisis with significant heterogeneity in risk profiles and outcomes. The national maternal mortality rate of 32.9 deaths per 100,000 live births in 2023 [6], is 2-3 times higher than other high-income countries and uniquely increasing [7]. In addition to mortality rates, 15% of women experience at least some pregnancy-related complication, with 10% experiencing maternal morbidity, and 3%(∼60,000) experiencing severe morbidity (life-threatening conditions) each year, such as eclampsia, postpartum hemorrhage, and cardiomyopathy [8, 9]. These statistics also mask profound disparities related to social determinants of health (SDoH): Black women face mortality rates of 50.3 per 100,000 live births, significantly higher than rates for White (14.5), Hispanic (12.4), and Asian (10.7) women. There also exist age-related disparities with women aged 40 and older experiencing 58.8 deaths per 100,000 live births compared to women 25-39 (18.1) and those less than 25 (12.5) [10, 19]. Other SDoH associated with these disparities in adverse outcomes include public or no insurance coverage, lower education levels, geographical location and political and cultural contexts [20], though there is limited literature and a dire need for further investigation into non-race and non-age SDoH that impact maternal health.

Despite over 80% of pregnancy-related deaths being preventable with early detection and intervention [21], our reliance on aggregate statistics limits our ability to identify individual risk profiles. This is further aggravated by under-diagnoses and under-reporting especially among socio-economic subgroups due to limitations in diagnostic tools, biases in health care and disparities in access to healthcare [22, 23]. Personalized risk prediction models are essential for targeted interventions across diverse populations, particularly for high-impact conditions. Hemorrhage, infection and cardiovascular/blood pressure related morbidities such as cardiomyopathy and preeclampsia contribute to the majority of deaths in the US and globally [11]. In the United States, postpartum depression is a factor in suicide-related maternal mortality, which contributes to 20% of maternal deaths in the postpartum period [12].

Although machine learning (ML) approaches have demonstrated promise for predicting maternal complications from EHRs, prior studies overwhelmingly rely on single-site and single modality data [15–18], limiting comprehensiveness and generalizability. Even though some studies such as Li et al. (2022) have attempted to rectify this, over 92% of their 108,557 pregnancies cohort were still drawn from one out of two hospitals in the Mount Sinai Health System to train models that predict preeclampsia [18]. These limitations raise concerns about model generalizability across diverse populations in healthcare settings, particularly given the documented socioeconomic disparities in maternal health. A major barrier has been the lack of a large-scale, diverse, multimodal, and multi-site maternal health dataset with sufficient depth to support both population-level characterization and robust ML-based risk prediction.

NIH’s “All of Us Research Program” (AoURP) unprecedentedly addresses this gap by presenting a United States-based, nationwide longitudinal cohort study with over 848,000 diverse participants and 470,000 associated electronic health records (EHRs) [1]. This enables development of robust predictive models from clinical sites across the United States, while examining generalizable correlations between EHR, socioeconomic factors, and pregnancy outcomes. Unlike traditional healthcare datasets which rely mainly on EHR, It also provides three additional rich modalities including wearables, genomics, and survey data, making it a uniquely powerful resource.

Although previous work on the *AoURP* have developed methods to robustly extract pregnancy episodes [24], or identified the potential applicabilities of the dataset in maternal health settings [33], no prior study has systematically evaluated its data quality and representativeness, characterized risk factors, or demonstrated its utility for training predictive ML models evaluated across the Social Determinant of Health (SDoH) surveys. Recent work leveraging wearable data for postpartum depression prediction [25] has shown promise but is limited by a very small pregnant-participant sample size (<60), underscoring the need for a scalable EHR-based approach as well as additional wearable data in the cohort.

We present the first comprehensive investigation of the pregnancy cohort in the *AoURP* dataset, which leverages longitudinal EHR data and SDoH Surveys to investigate and develop predictive ML models on seven critical adverse outcomes: Depression and Anxiety, Gestational Diabetes, Preterm Labor, Preeclampsia, Miscarriage, Cardiomyopathy, and Eclampsia. Our objectives are to determine whether ML models can predict maternal health outcomes using the *AoURP* dataset, determine whether novel risk factors are identified, and demonstrate the utility of the pregnancy cohort extracted from AoURP for ML research, delineating its strengths and limitations.

Our primary contributions are:

1. Curation of reproducible preprocessing pipelines, which includes pregnancy episode and outcome identification, multi-source data extraction, and feature harmonization across a nationally diverse population. The code and cleaned dataset is directly available on the All of Us research platform for use by registered users.
2. Development of state-of-the-art predictive machine learning models across different maternal morbidities: miscarriage, gestational diabetes, preeclampsia, and cardiomyopathy, establishing performance benchmarks for future AoURP-based maternal health studies.
3. Identification of potentially novel risk factors/features not currently used in standard clinical practice, which may warrant additional research.
4. ML model fairness assessment using SDoH metrics derived from survey data to understand how ML models perform across subgroups and, providing an initial assessment of equity considerations for ML models within this nation-widecohort.

## Results

### Cohort selection, data filtering and feature extraction yields heterogenous data associated with social determinants of health

Through cohort identification and preprocessing methods, we constructed the pregnancy dataset consisting of 20,253 subjects and 27,525 distinct pregnancy episodes (Fig.1a, Fig.1b left). Seven adverse outcome labels were extracted from the different indicators: Preterm Labor, Depression and Anxiety, Gestational Diabetes, Pre-eclampsia, Miscarriage, Cardiomyopathy, and Eclampsia. These were selected based on relative impact on maternal mortality and morbidity, and actionability if identified early. We extracted EHR features from the top 1000 occurring conditions, labs and measurements in the EHR domain, and applied extensive data cleaning, anomaly detection, and unification methods to construct a strong and comprehensive pregnancy dataset (Fig.1b center). As illustrated in Fig. 1d, patient timelines were aligned to individual pregnancy episodes, where a data point was included if it was recorded within 280 days prior to the end of pregnancy, and up to a predetermined prediction gap (defaulted to be 28 days) before an adverse outcome onset or a normal delivery.

**Figure 1:**
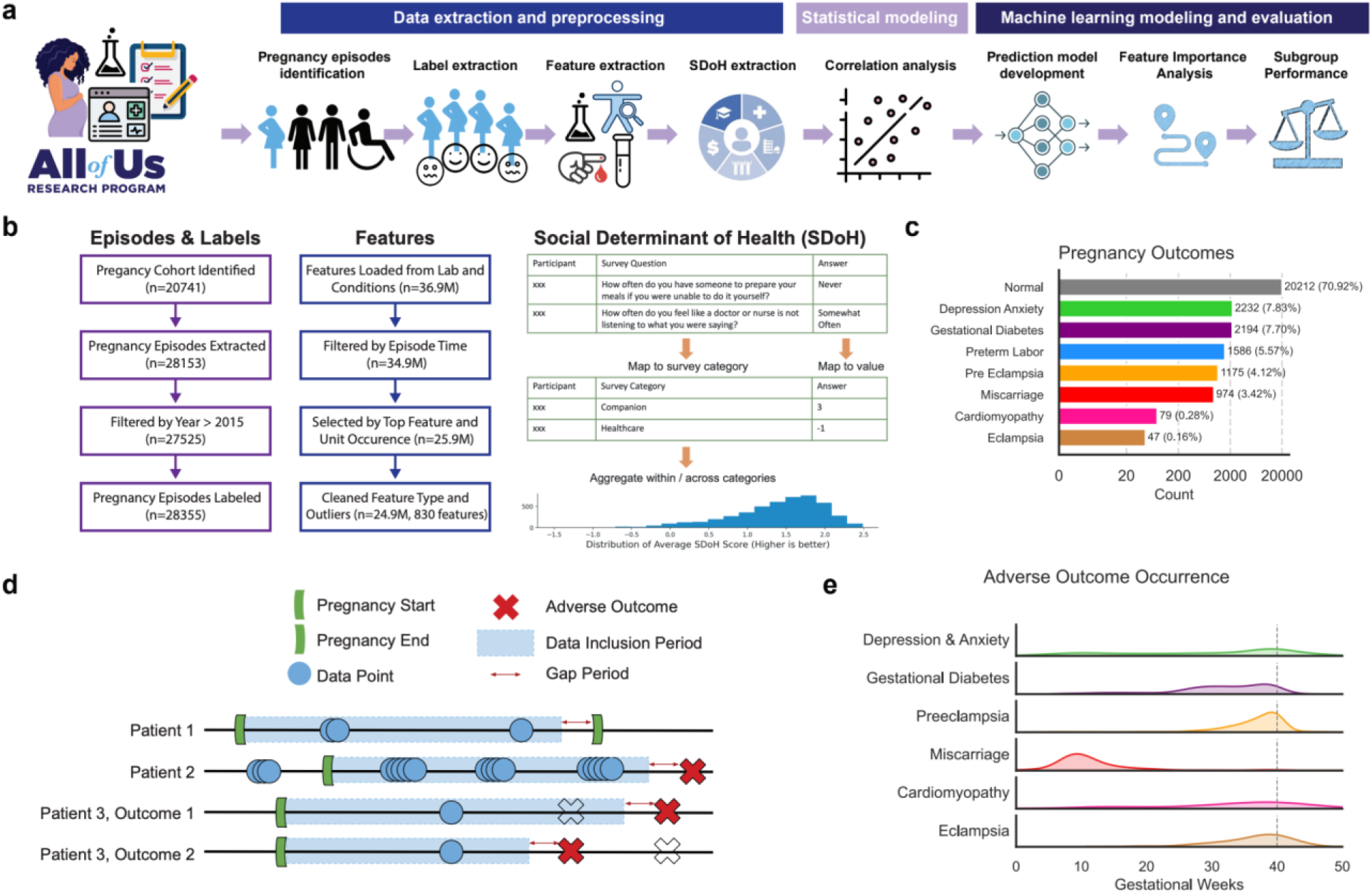
Methodology overview: Data preprocessing, validity analysis, and ML pipeline. a) End-to-end study workflow. b) Flowcharts summarizing the construction of *AoURP* maternal health cohort, extracting pregnancy episodes, features, and SDoH from the dataset, c) Prevalence of the seven adverse pregnancy outcomes used for prediction tasks. d) Examples of sample construction from patient timelines. e) Onset timing for each adverse maternal event.

Additionally, we identified seven SDoH features from the surveys by defining and categorizing questions and mapping answers to continuous values (Fig.1b right, Appendix B). We defined 7 SDoH categories: Companion, Discrimination, Efficacy, Healthcare, Neighborhood, Religion, and All SDoH (an average of all previous categories). Greater than 98% of participants took the basic demographics survey (table 1) which was used for age and race-related subgroup fairness evaluations. However, only 21% of participants had SDoH survey results and were included in the SDoH sub-group fairness evaluations.

**Table 1.** Subject Basic Characteristics Summary*.

| Variable | Counts (Percentage%) or Median (Interquartile Range) |
| --- | --- |
| Participants | 20,253 |
| Pregnancy Episodes | 27,525 |
| Age | 29.22 (24.63, 33.64) |
| **Race \*\*** |  |
| None Indicated | 7479 (36.93%) |
| White | 7411 (36.59%) |
| Black or African American | 3040 (15.01%) |
| Other | 1699 (8.24%) |
| Asian | 654 (3.23%) |
| **Education** |  |
| College | 12272 (60.59%) |
| High School | 5406 (26.69%) |
| Others | 2195 (10.84%) |
| Unknown | 380 (1.88%) |
| **Smoking** |  |
| Never/Rare (< 100 cigarettes) | 15243 (75.65%) |
| Often (>= 100 cigarettes) | 4379 (21.73%) |
| Unknown | 527 (2.62%) |
| **Geographic Region** |  |
| Northeast | 8012 (39.56%) |
| West | 6789 (33.52%) |
| Midwest | 2898 (14.31%) |
| South | 1888 (9.32%) |
| Unknown | 666 (3.29%) |

\*Counts may not add up to the participant number since less than 2% of the participants did not complete “The Basics” survey. \*\*Race was extracted from “The Basics” survey of the All of Us dataset. In the original question, race and ethnicity information were asked in the form of a single multi-choice question [13], then transformed to two columns “race” and “ethnicity” based on comprehensive transformation rules [14]. In this study, we use only the derived “race” variable. It is worth noting that the majority of individuals categorized as “None Indicated” in the race variable self-identified as Hispanic or Latino in the ethnicity field. “Other” category includes “More than one population”, “Another single population”, “None of these”, “PMI: Skip”, and “I prefer not to answer”.

We verified the soundness of our curated dataset focusing on the adverse outcomes extracted (Fig.1c, Fig.1e). The epidemiological prevalence of the seven adverse outcomes matched the literature [34, 35, 37, 38, 39, 40]. The onset time of the outcomes relative to the pregnancy duration also matched clinical expectations. For example, miscarriage occurrence matched the definition of the loss of pregnancy in the first 20 weeks [38]. Depression and anxiety distributed more equally throughout the peripatrum period [40], compared to other adverse outcomes which were centered around the end of pregnancy.

### Data cleaning improves usability but demonstrates challenges with multi-institute datasets

We considered additional data quality measurements including unit unification, collapsible features, and the degree of missingness. Even though features between hospitals were somewhat unified by common systems such as ICD code, there were high levels of inconsistencies observed in the units of lab measurements, which is a fundamental challenge in any multi-hospital dataset. After unit cleaning, the percentage of inconsistent units decreased from 24% to 0% (Appendix C). Another source of data inconsistency was the use of multiple feature names for the same concept, caused by different measurement methods included in the feature name. By conducting N-grams analysis and consulting with physicians, we identified 164 potentially collapsible features within six concept groups (Appendix D), and collapsed them to 85 features. Finally, we assessed the degree of missingness of the features. We found that among the 1,000 extracted features, 64% occurred in only fewer than 1% of pregnancy episodes. This outlined another challenge in using a large EHR-based dataset with comprehensive features. Outside of common lab tests such as complete blood count and vital measures, the other lab tests are much less ordered and diagnostics codes are also much more sparse. If formulated in a tabular manner, the majority of the columns will consist of missing values, which motivates us to consider sequential data formulation methods.

### Statistical correlations identify features associated adverse outcomes

We used multiple logistic regression models adjusted for age, race, and smoking status to determine correlations between each feature extracted from both the EHR data and survey, and the adverse maternal outcomes, corrected for multiple comparisons. The sequential features were engineered to extract the mean, median, max and min over the period of interest (start of pregnancy until gap days of 28). Negative Log of p-values from statistical correlations for features and outcomes are shown in Fig. 2 with the highest correlated features labeled for each outcome. Expectedly, we found several clinically expected correlations: blood pressure was correlated with preeclampsia, a form of hypertensive disorder, and glucose challenge tests were strongly correlated with gestational diabetes. However, we also found a few clinically under-used but logical associations, such as cholesterol associated with miscarriage, which prompted further analysis.

**Figure 2:**
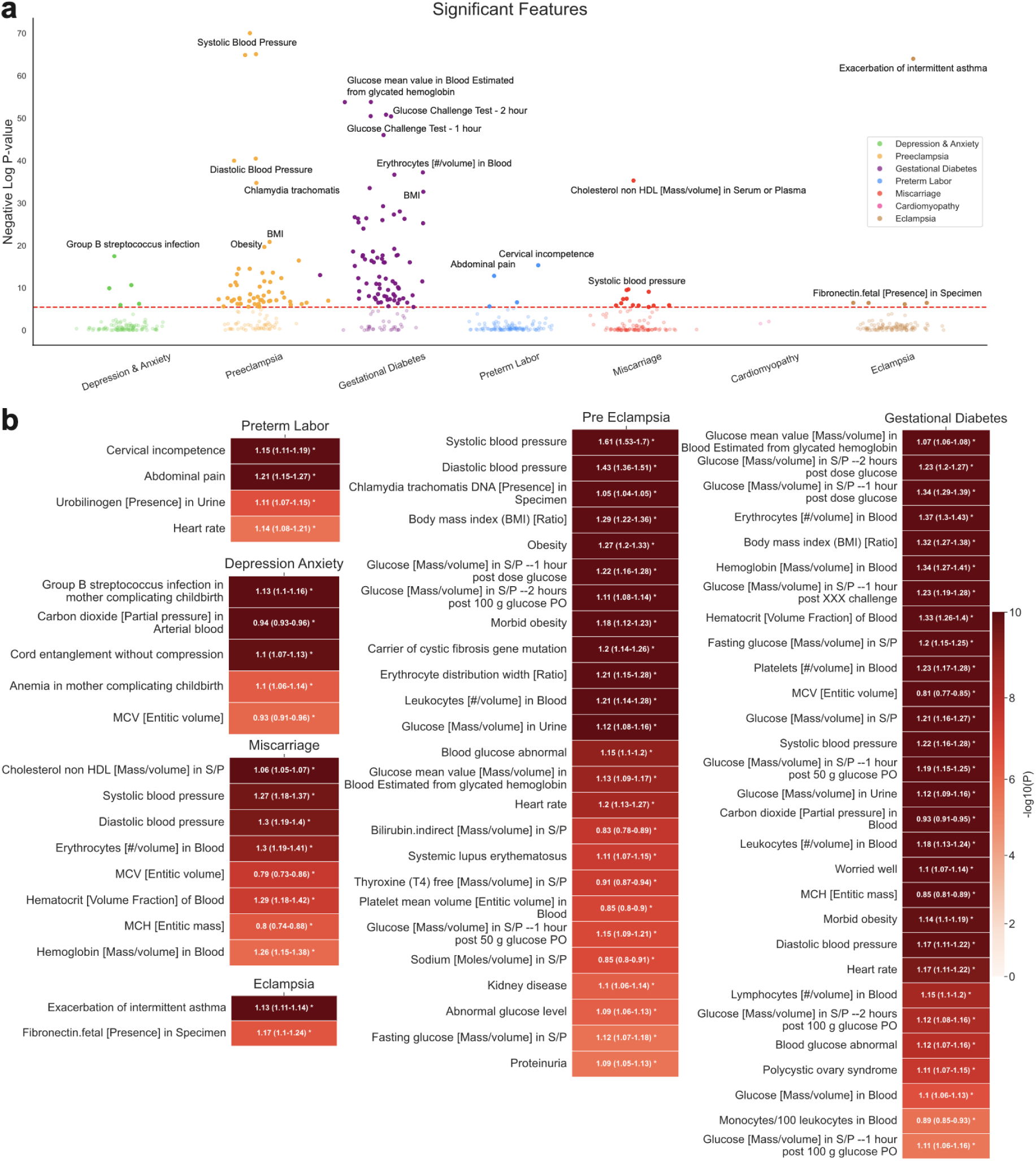
Statistical correlations of features with adverse outcomes. a) Plot of -log10(p) values on the y-axis for all features per outcome, grouped by each adverse outcome. Features above the dashed red line (Bonferroni Threshold) are considered statistically significant after correction for multiple comparisons. b) Heatmap summarizing statistically significant feature– outcome associations. Red color intensity of the boxes represents −log10(p) values, while overlaid white text reports odds ratios (ORs) with corresponding confidence intervals for each adverse outcome.

### ML models enable prediction of adverse pregnancy outcomes from the dataset

Machine learning models (logistic regression, XGBoost, and LSTM) were trained on the data to predict adverse outcomes 28 days in advance, notably achieved an AUROC of 0.923 for miscarriage, 0.751 for gestational diabetes, 0.748 for preeclampsia, and 0.761 for cardiomyopathy with corresponding F1 scores of 0.686, 0.587, 0.521, and 0.579. Compared to logistic regression and XGBoost, LSTM performed the best for all outcomes except for preeclampsia where XGBoost performed the best (AUROC=0.748). We show the detailed performance (mean AUROC and mean macro-averaged F1 scores) for all models and outcomes (table 2 and Fig 3a), the ROC and the precision recall curve (Fig 3b,c) for XGBoost models and LSTM models (Fig 3e,f). Additionally, we performed analysis on the effect of changing gap days (defined as the data cutoff time prior to adverse outcome onset), on the performance of the model. Performance generally decreased as gap days increased from 2 weeks to 8 weeks (Fig 3d,g). In all remaining analysis in the paper, we focused on using the results from 28 gap days, which indicates the models’ predictive power one month in advance of the adverse outcome onset, enabling sufficient time for clinical actionability.

**Figure 3:**
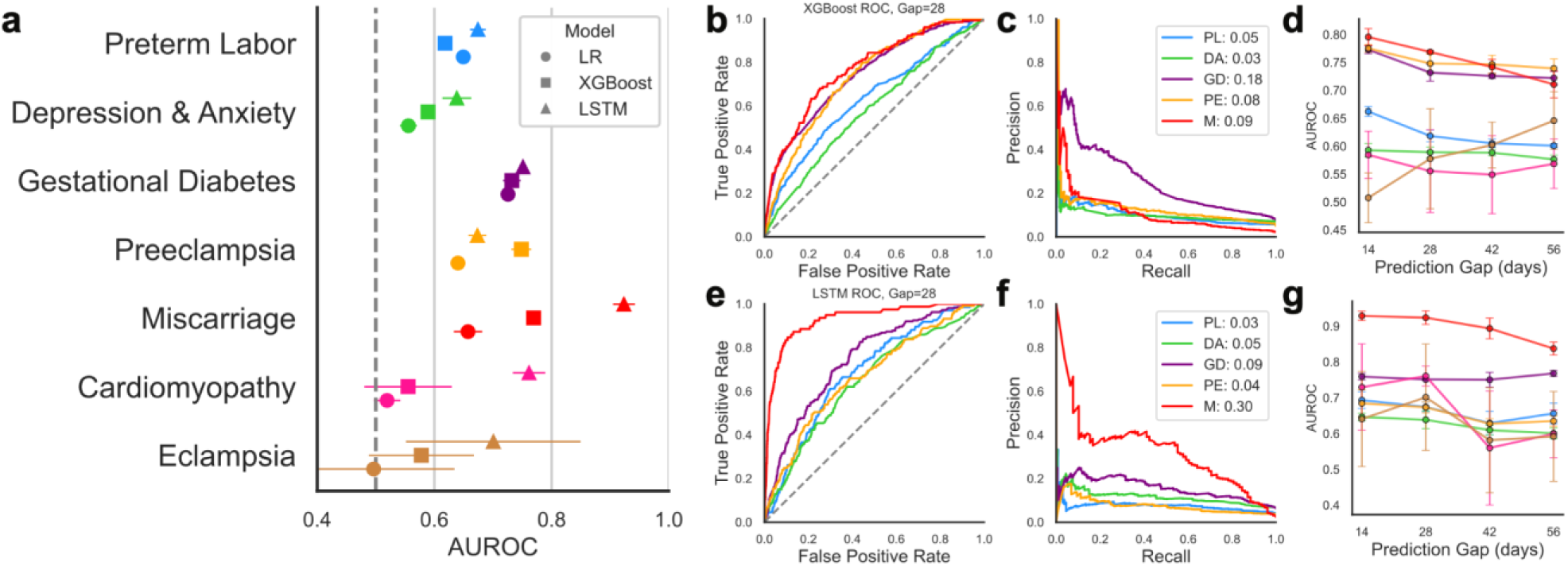
(a) AUROC comparisons for all models, across all outcomes. Error bars are standard deviation from 4-fold cross-validation performance. (b) ROC curves for XGBoost models in the first fold of training for each outcome. (c) Precision-recall curves for XGBoost models in the first fold of training for each outcome. The AUPRC values are shown in the legend. Outcome names are abbreviated: (PL=Preterm Labor, DA=Depression & Anxiety, GD=Gestational Diabetes, PE=Preeclampsia, M=Miscarriage). (d) Gap days analysis for XGBoost models (e,f,g) Corresponding plots for LSTM models.

**Table 2.** ML Models Performance Summary.

|  | Cardiomyopathy |  | Depression and Anxiety |  | Eclampsia |  | Gestational Diabetes |  | Miscarriage |  | Preeclampsia |  | Preterm Labor |  |
| --- | --- | --- | --- | --- | --- | --- | --- | --- | --- | --- | --- | --- | --- | --- |
| Model | AUROC | F1 | AUROC | F1 | AUROC | F1 | AUROC | F1 | AUROC | F1 | AUROC | F1 | AUROC | F1 |
| LR | 0.519 | 0.516 | 0.556 | 0.525 | 0.496 | 0.507 | 0.725 | 0.597 | 0.657 | 0.528 | 0.64 | 0.569 | 0.649 | 0.572 |
| XGB | 0.555 | 0.503 | 0.589 | 0.535 | 0.578 | 0.5 | 0.732 | 0.605 | 0.769 | 0.574 | **0.748** | **0.579** | 0.618 | 0.546 |
| LSTM | **0.761** | **0.521** | **0.638** | **0.553** | **0.700** | **0.507** | **0.751** | **0.587** | **0.923** | **0.686** | 0.673 | 0.549 | **0.674** | **0.537** |

\* LR=logistic regression, XGB = XGBoost.

We conducted interpretability analysis for the ML models which have sufficient positive label supports. SHAP values were used for XGBoost feature importance (Fig. 4a) and leave-one-out feature ablation method was used for LSTM model feature importance (Fig. 4b). We found that XGBoost’s feature importance was positively associated with the odds ratio determined by correlation analysis (Fig. 4c, appendix J). Feature importance of the LSTM was in turn generally positively associated with the availability of the feature (Fig. 4d, appendix K).

**Figure 4:**
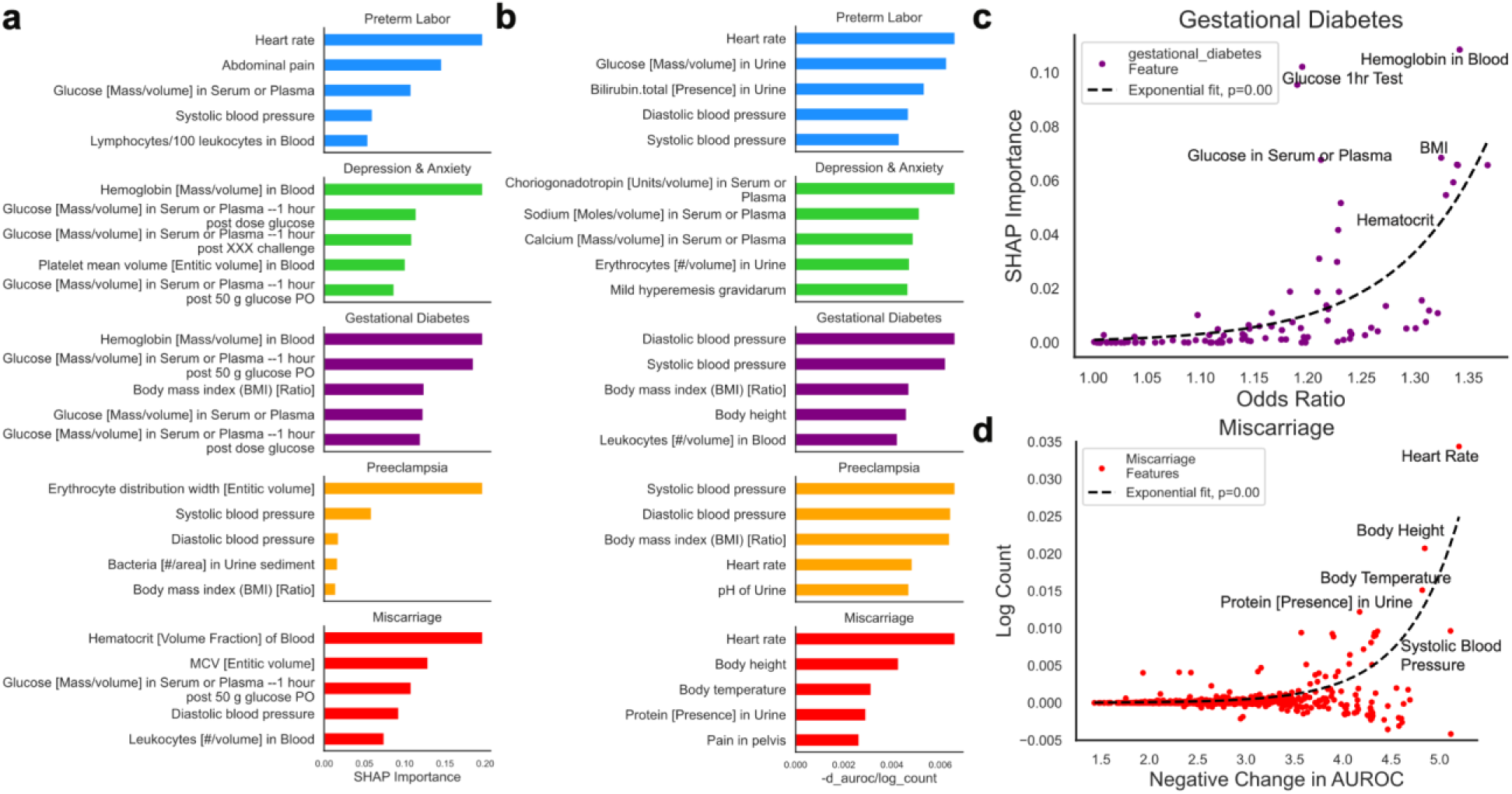
(a) Top 5 features with highest SHAP importance values from trained XGBoost models. (b) Top 5 features with highest drop in performance when removed from trained LSTM models, log normalized by number of occurrences. (c) There exists an exponential relationship for trained XGBoost models between odds ratio and feature importance (p=1.53e-8, two-sided t test on growth parameter). Example of outcome gestational diabetes shown. (d) When LSTM is trained to predict miscarriage, there exists an exponential relationship between feature importance and feature occurrence (p<1e-10). Example of miscarriage outcome shown.

For each outcome, we extracted two sets of non-mutually exclusive notable features: one set from statistical correlation analysis and another set from ML interpretability analysis. Features derived from correlation more frequently exhibit large differences between negative and positive groups, such as glucose for gestational diabetes. Some features showed no significant difference in mean or median, but significant difference in tail distributions, for example glucose in urine for several adverse outcomes. Some ML-based notable features exhibited no observable difference. For instance, when broadly looking at the population level, heart rate showed minimal difference between the negative (85.3 (74.7-96.0)) and positive group 84.7 (73.3-96.2) despite being ranked a highly important feature for model prediction, a result of the complex, non-linear, dynamic approach of ML models.

### Physician ratings reveal unexpected notable features associated with adverse outcomes

Four physicians (obstetricians and family medicine practitioners) evaluated the set of notable features across two metrics:

1. **Expectedness:** To what extent they expect the features to be associated with the adverse outcome (expectedness)
2. **Use in clinical practice**: To what extent the features are currently considered as a risk factor in clinical practice.

The ratings from the four physicians showed fair inter-rater agreement with Kendall’s W of 0.535. We found that the majority of the features were rated highly by the physicians for expectedness and frequently considered in clinical practice (Appendix G), for example glucose-related features and gestational diabetes (Fig. 5a). A few notable features were rated as unexpected and under-considered in the clinical practices. Complete Blood Count (CBC) was one of the most prevalent labs generally available in this dataset. Seven CBC-derived features (erythrocytes, hematocrit, hemoglobin, MCH, MCV, erythrocyte distribution width, platelets, and leukocytes) demonstrated strong associations with gestational diabetes and miscarriage, which clinicians rated as largely unexpected (Fig. 5b,c). Carriers of the cystic fibrosis gene mutation were 62% more likely to develop preeclampsia, yet clinicians rated it as both unexpected and rarely considered in current practice (Fig. 5d). Overall, these features tend to have larger disagreements and variance.

**Figure 5.**
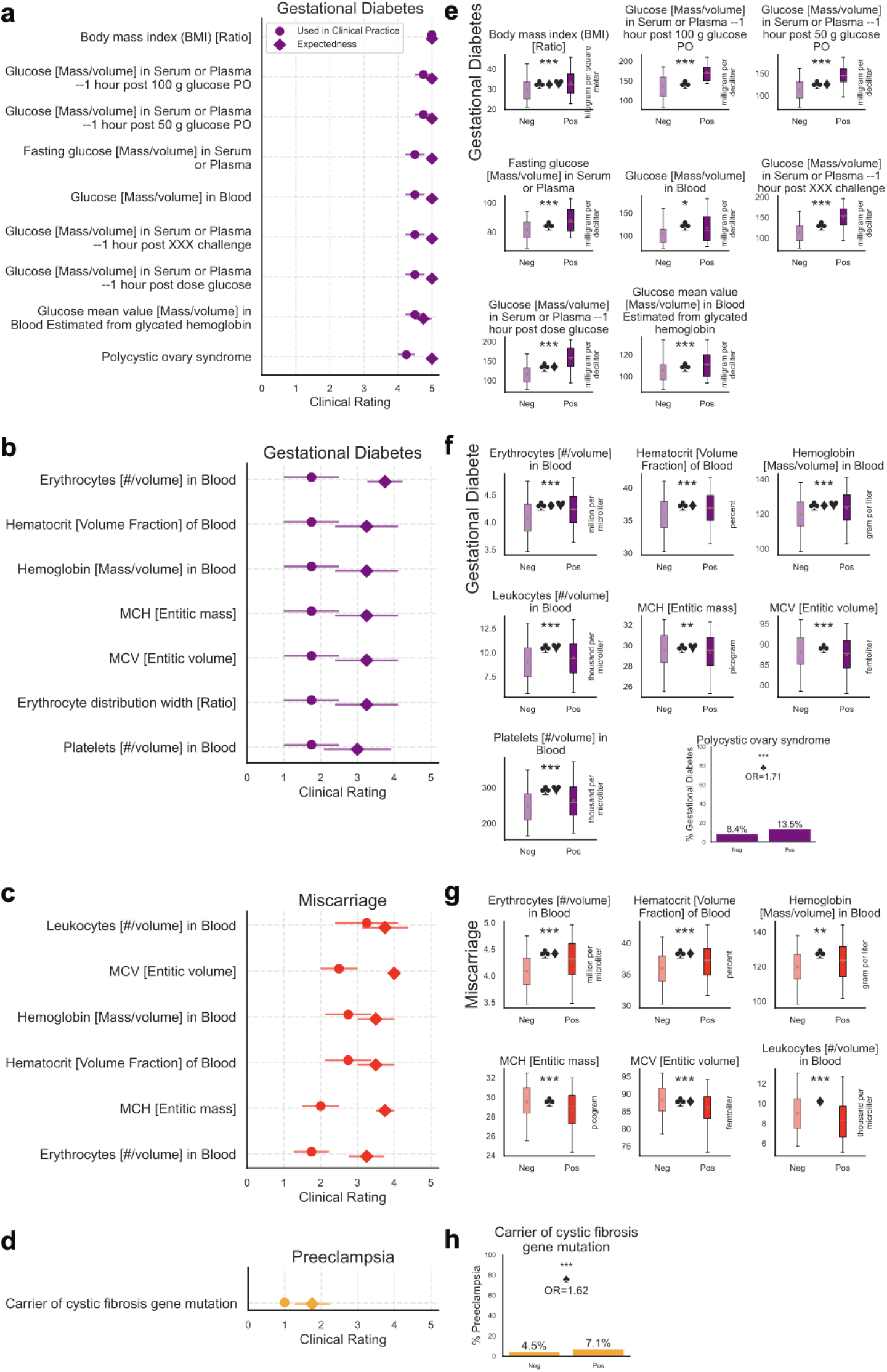
Examples of Clinical values of the identified notable features. **a-d:** Mean clinician ratings (n=4) on a 5-point scale for the feature associations with outcome. Clinicians rated the expectedness of each association (circle) and the extent to which the feature is considered as a risk factor for the adverse outcome in current practice (diamond). **e-h:** Direct Group Visualizations. Continuous features are shown using box plot (5%, 25%, 50%, 75%, 95% percentile, gray scatter point indicates mean, p-value indicated as *(p<0.01),**(p<0.001),***(p<0.0001), or n.s. (no significance). Discrete features’ percentages are shown. Symbols indicate feature sources (can have multiple): ♣ (from correlation), ♦ (from XGBoost), ♥ (from LSTM). Complete versions of the visualizations are provided in Appendices G and H.

### Fairness evaluation identifies SDoH factors statistically correlated with adverse outcomes, but reveals no significant disparities in ML performance across SDoH subgroups

Depression and anxiety was the only adverse outcome that was significantly associated with the SDoH features (Efficacy, Neighborhood, All SDoH). Efficacy, defined as one’s belief and confidence in themselves having control over their lives, showed a significantly negative correlation with depression (odds ratio = 0.71) meaning low efficacy could be a strong risk factor for depression and anxiety (Fig. 6a). Detailed questions and categorizations are provided in appendix B.

**Figure 6:**
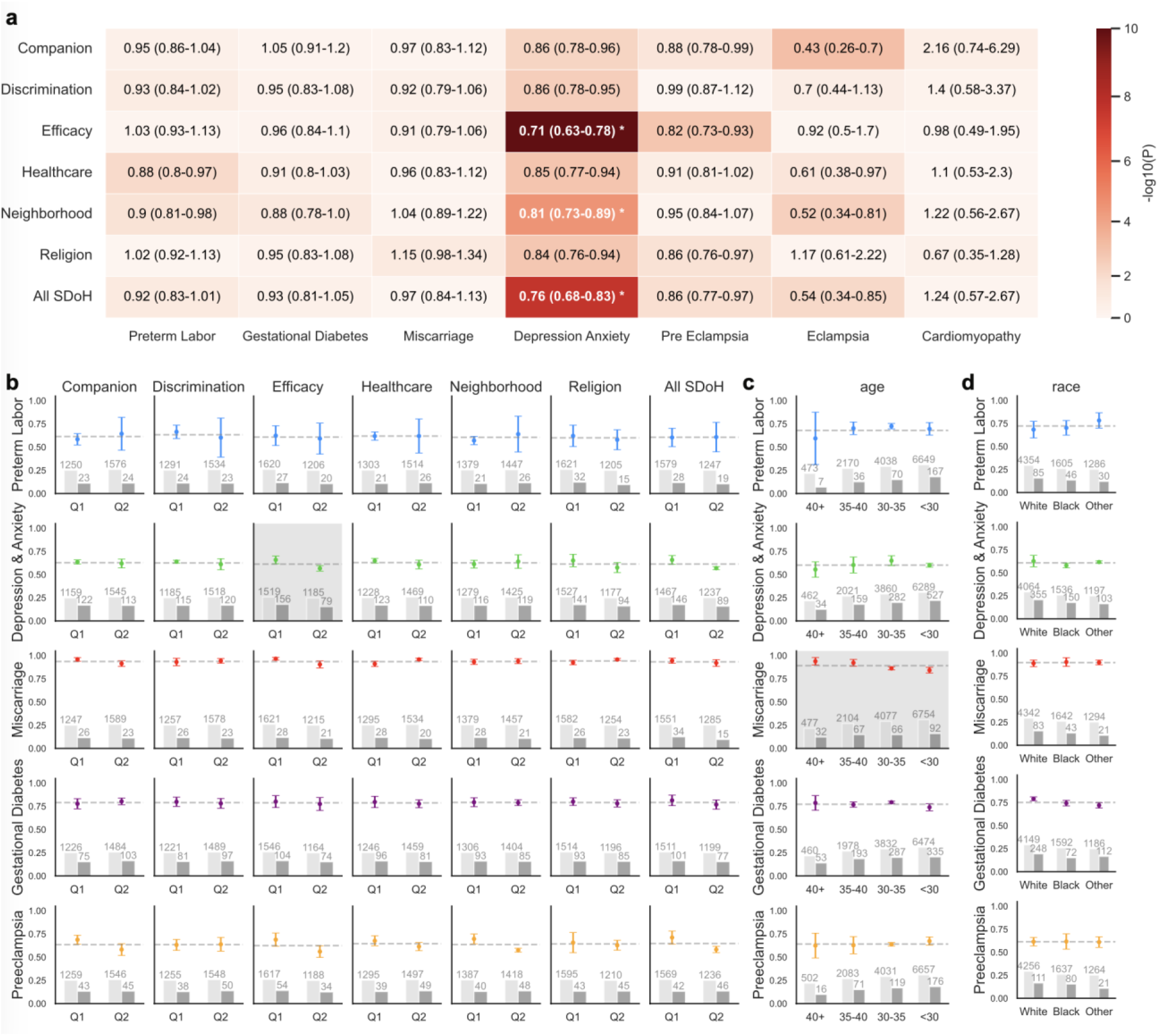
Fairness evaluations. (a) Direct correlation of the SDoH features with each outcome. Color indicates -log10(p) value, and overlaid numbers indicate OR with confidence intervals similar to Fig. 2b. (b-d) Subgroup performance analysis of machine learning models stratified by SDoH status (b), age (c), and race (d). For SDoH analysis, Q1 denotes individuals in the lower 50% (worse SDoH status) and Q2 denotes the upper 50%. Points indicate mean AUROC across four-fold cross-validation and error bars indicate variability. Vertical bars and counts at the bottom indicate the number of negative labels (light gray) and positive labels (dark gray) in each subgroup.

Due to concerns about bias in ML models, we evaluated the fairness of trained models by assessing subgroup-specific performance. We divide participants into three categories of subgroups including age, race, and survey-reported SDoH. Note that the SDoH subgroups were also included as input features as a bias mitigation technique during the training of machine learning models. We then evaluated if the models perform consistently for different subgroups.

We found no significant unfairness in our ML models across SDoH subgroups, there were two marginally significant cases that warrants attention. First, as it was also revealed in correlation, the model performed unfairly when two efficacy groups were considered (Fig. 6a, row2, col 3, highlighted in gray). We observed decreasing model performance with decreasing age (Fig. 6c, row 3, highlighted in gray). however this was not significant. Note that we did not include age as a feature during predictive model training.

## Discussion

This study presents the first in-depth exploration utilizing derived longitudinal and multimodal EHR and survey features from the *AoURP* dataset for development of machine learning models for predicting maternal morbidities. Our analysis focused on a large, heterogeneous, multi-site US population, demonstrating the feasibility of training high-performing ML models on this unprecedented type of real-world data and evaluating model fairness based on SDoH survey data.

We trained and evaluated three classes of ML models: logistic regression, decision trees-based XGBoost and a time-series deep learning based LSTM neural network., XGBoost and LSTM yielded the best performance, with XGBoost performing best for preeclampsia and LSTM performing best for predicting all the other adverse outcomes. These differences in performance likely reflect the types of features most informative for each outcome. Deep learning based time series models can capture nuanced sequential patterns, and thus better utilize features collected frequently during pregnancy such as heart rate and blood pressure. LSTM importance scores were positively associated with the number of measurements per feature. In contrast, tree-based XGBoost models which excel on tabular data, performed better in cases where features had strong aggregated correlations with outcomes such as glucose tests. Together, these results suggest that the maternal morbidity outcomes differ in their underlying modeling complexity. Outcomes such as miscarriage may require models capable of capturing higher-order temporal interactions, whereas more pathophysiologically well-characterized conditions such as gestational diabetes may benefit from simpler, more interpretable modeling approaches.

Our best performing model was for miscarriage prediction, achieving an AUROC of 0.923, with important features including high-frequency features like blood pressure and heart rate. This represents state-of-the-art performance for predicting miscarriage onset from routine EHR data. Prior work based on ultrasound data reported an AUROC of 0.857 [31], underscoring the potential value of large-scale, longitudinal EHR data for studying miscarriage risk. Unfortunately, miscarriage is not a clinically preventable outcome, beyond provision of palliative care There is a need for further studies to carefully consider the clinical and ethical implications of these ML models, and how they can benefit patients and impact care.

For preterm labor, our best model achieved an AUROC of 0.674, in line with published studies. Huang et al. (2024) achieved AUROC scores of 0.62-0.71 using Elastic net regularized logistic regression models with data from a multi-center nuMoM2b [27]. Abraham et al. (2022) achieved AUROC scores of 0.63-0.72 using an XGBoost trained on data from Vanderbilt Health System and verified on data from the University of California, San Francisco Health System [28].

For other outcomes, to the best of our knowledge, no prior work has reported multi-site prediction performance. The AUROC of our preeclampsia prediction model (0.748) is lower than the AUROC of 0.82 (intrapatrum) reported by Li et al (2022) from models trained on EHR data from Mount Sinai Hospital System [18], or AUROC of 0.955 reported by Li et al (2021) from models trained on a single hospital in Shanghai [26]. Similar to the latter study’s findings, our study also found that for predicting preeclampsia, XGBoost was the best performing model and, glucose, blood pressure, and obesity were the most predictive features [26]. Our models predicted eclampsia at 0.700 AUROC, and did not conduct further analysis due to cohort size limitations. We found no other studies that predict eclampsia from EHR.

The best AUROC for predicting depression and anxiety was 0.638, lower than the best AUROC of 0.72-0.74 reported by Amit et al. (2021) which applied XGBoost to EHR data from a UK cohort of 266,544 women with 35,708 postpartum depression cases [29]. Our gestational diabetes model performance of 0.751 was lower than the AUROC of 0.85 from a large cohort study in Israel of 588,622 women (Artzi et al. 2020) [30]. We achieved an AUROC of 0.761 on cardiomyopathy but did not perform further analysis due to limited sample size, however existing literature on large scale studies have found AUROC of 0.80 using logistic regression (Davis et al. 2021) [32]. Our models’ performance were generally lower than studies that used single-hospital cohorts, suggesting that while using multi-site, nation-wide EHR dataset is a better representation of the real-world diversity, meanwhile also more challenging.

Most features we identified through statistical analysis and through ML feature importance analysis aligned with physician clinical use and expectedness. However, others such as associations between cystic fibrosis gene mutation and preeclampsia, as well as between CBC, gestation diabetes and miscarriage, were rated as unexpected and not previously emphasized in standard clinical decisions (Fig. 5b-d). Although cystic fibrosis carrier mutations are among the most common gene variants, with an onset rate of approximately 1 in 2,500 in Caucasian populations and an estimated >10 million carriers in the United States [41, 42], their implications for maternal health have received limited attention. While recent literature has highlighted broader disease risks associated with these mutations [42], their potential contribution to adverse pregnancy outcomes remains underexplored. Recent work has also demonstrated the predictive value of routine CBC tests in disease forecasting [43], and our results further support the potential of these features in identifying gestational diabetes and miscarriage risk. Notably, abnormalities across multiple CBC measures may carry meaningful signals, even when such patterns are not readily apparent from population-level group statistics. This highlights the value of individual-level thresholds for common laboratory markers in clinical risk stratification [43]. Future work should investigate these connections more systematically both in the All of Us dataset and external cohorts.

A limitation in our study that may account for underperformance is our unified prediction framework across all adverse outcomes. While this approach streamlines data preprocessing and facilitates generalizable modeling, it potentially comes at the cost of overlooking outcome-specific nuances that tailored feature engineering or specialized models might capture more effectively. However we note that our performance is within range of most currently published prediction methods despite being a larger, more diverse dataset. Future work could focus on developing optimized, outcome-specific modeling strategies using our curated dataset, modeling pipeline and identified features.

Another limitation of the study was that the population was limited to the US, and replicating this work in other regions with high maternal morbidity risk is challenging due to data limitations. However, we identify simple features that are routinely measured during pregnancy which may enable global replication. An additional limitation is the high data imbalances between SDoH subgroups, and underrepresentation of some racial groups, requiring further data collection for members of these groups.

While the ML models demonstrate promise for predicting maternal morbidity outcomes, and identifying potential features not currently used for clinical decision making, this study only provides an initial baseline retrospective analysis which requires further clinical validation. Prospective clinical validations, with additional stakeholder input, would greatly benefit demonstration of robustness, generalizability, and potential for clinical implementation. Finally, given the higher risk for worse maternal health outcomes among underrepresented SDoH groups, ML models need to be continuously evaluated through additional data collection and improved fairness mitigation methods.

## Methods

### Dataset Description

The All of Us Dataset is an NIH-funded biomedical dataset incorporating EHR, genomic, survey, and wearable data with over 872,000 diverse participants in the U.S. The study uses the AoURP registered tier v8 dataset, and all analyses were conducted on the researcher workbench cloud platform. The EHR was collected from hospitals across the US. Participants, and a subset of them also filled out SDoH surveys which were linked to the EHRs. Participants were provided informed consent under a human participant research protocol reviewed by the institutional review board of the AoURP (protocol 2021-02-TN-001).

### Cohort selection

In this study, we aimed to focus on the pregnant cohort and use data from three categories: Conditions (Medical concepts that describe the health status of an individual, e.g. Abdominal Pain), Labs & Measurements (Medical concepts that capture values resulting from examinations or tests, e.g. Diastolic Blood Pressure) from EHR, and Social Determinants of Health from Survey (How common is vandalism in your neighborhood?).

We adapted existing approaches for pregnancy episode extraction from All of Us Dataset [24], but introduced substantial modifications to accommodate our adverse outcome labeling and ML pipeline. We first identified the pregnancy cohort of interest and the corresponding pregnancy date range (i.e. pregnancy start date and pregnancy end date). Within the data, there were two main categories of conditions that could both indicate pregnancy as well as the specific duration of pregnancy: delivery findings, that either indicate delivery with or without complications (e.g. ‘Mother delivered’), and gestational findings (e.g. ‘Gestation period, 32 weeks’). For the former type of information, it was assumed that they indicated the precise end time point of pregnancy. For the latter, we inferred the pregnancy start date by gestational weeks, then calculated the pregnancy end date assuming a fixed duration of 280 days to facilitate standardized machine learning modeling. Although this approximation for gestation information simplified the natural variation in human pregnancy length [2], it did not compromise the study’s core findings and was further mitigated by the median-aggregation approach described below. Since the length of preterm labor episodes were inherently shorter, this assumption may introduce additional prepartum data, which we acknowledge as a limitation.

Building on the cohort where at least one of the temporal-indicating conditions were present, we then solved two inconsistency problems. Firstly, one participant could have multiple pregnancy episodes. The data itself did not distinguish between the episodes, so we grouped pregnancy-duration findings according to the criteria that if two conditions were separated by at least 365 days, then they were counted as two different episodes. Secondly, within one episode, oftentimes multiple duration-identifying conditions were available and the pregnancy end date had inconsistencies of five days as determined by different features, likely due to the approximation of gestational findings which had only the nearest week. We alleviated this by taking the median of all the available pregnancy end days, yielding a robust date.

This yielded a cohort of 20,253 subjects who had 27,525 distinct pregnancy episodes (table 1) with pregnancy episodes filtered to fall in the range of 2015 to 2023 (Appendix E).

### Extracting and pre-processing outcome labels

We selected a total of seven maternal health adverse outcomes that have substantial impacts on preventable maternal morbidity and mortality rates, and are potentially also actionable [11,12], as the labels of our classification problem: Preterm Labor, Depression and Anxiety, Gestational Diabetes, Pre-eclampsia, Miscarriage, Cardiomyopathy, and Eclampsia. For each adverse outcome, there were multiple indicator conditions in the data, so they were labeled one of the above 7 classes (Appendix A).

We matched each outcome label to their corresponding pregnancy episodes according to their onset time. There were rare circumstances where one pregnancy episode had more than one adverse outcome. In that case we treated them as positive labels in more than one adverse outcome classification accordingly. Subsequently, we marked all episodes without adverse outcomes as Normal.

### Extracting and pre-processing EHR features

Features and values were pulled from two sources in the All of Us Dataset: a) Conditions, which contained occurrence of diagnosis, patient-reported symptoms and findings, and b) Labs & Measurements. Based on the cohort defined earlier, the top 1000 most frequently occurring features were retrieved and used for downstream ML tasks. Note that the condition codes already used in either cohort selection or outcomes labels were excluded.

Being sourced from multiple databases, the All of Us data was highly heterogeneous and very frequently contained missing values and malformed data. We executed extensive data cleaning steps to increase the signal to noise ratio.

We first conducted an n-gram search to identify similar or identical features. We collapsed them into a single feature based on review by an Obstetrician (Appendix D). For example, Basophils/100 leukocytes in Blood by Automated count’ and ‘Basophils/100 leukocytes in Blood by Manual count’ are combined as one feature since they only differ in measurement method.

We then address the unit inconsistency issue for the continuous features. Individual laboratory measurements were frequently recorded in multiple units, sometimes exceeding 20 variants for a single feature, likely reflecting the difference in guidelines and measurement equipment between hospitals. On average, the most dominant unit constituted 76% of all the entries of the features. We therefore retained measurements expressed in the dominant unit and discarded entries recorded in alternative units. For example, ‘Glucose [Mass/volume] in Serum or Plasma --1 hour post dose glucose’ had 6 different units presented in data: ‘milligram per deciliter’, ‘No matching concept’, None, ‘microgram per deciliter’, ‘milligram per milliliter’, ‘mg/dL’. Only the data entries with the first dominant one “milligram per deciliter” was taken and the rest were dropped. Categorical values were additionally differentiated and one-hot encoded.

We show examples of synthetic Patient Timelines and Data Inclusion in Fig. 1d. Features were either collected from *Labs & Measurements* or *Condition*. The inclusion window of data was within 280 days prior and up to a certain amount of gap day, defined as the number of days prior to adverse outcome onset during which observations were excluded to prevent information leakage (Fig. 1d). One patient could have multiple adverse outcomes, as shown for Fig. 1d, patient 3. In this case, they were treated separately as two positive episodes in two adverse different outcomes shown by two overlapping blue boxes. Data availability varied significantly between patients: for example, in Fig 1d, patient 2 has considerably more data than patient 1 or 3.

### SDoH surveys preprocessing and subgroup ranking

All of the Us dataset includes surveys incorporating various fields that a subset of participants answered. In this study, we are particularly interested in the social determinants of health (SDOH) extracted from the survey, which are designed to reflect the participant’s social and economical status, alongside other more common demographic info like race and age. The surveys are collected in the date range from November 2020 to June 2022, with the median month of July 2021, which generally overlaps with the majority of pregnancy episodes dates in the cohort selection mentioned above.

We used the social determinant of health section of the survey and transformed the qualitative answers to quantitative measures by hand labeling each answer to continuous numbers (e.g. Strongly disagree maps to −3 and somewhat agree maps to 1, appendix B). We also grouped all questions based on their focus on one specific SDOH category: Companion, Discrimination, Efficacy, Healthcare, Neighborhood, Religion, and finally All, where “All” is an average to all previous categories to indicate overall SDoH (Appendix B). The scores of answers are then averaged within the categories per subject, and a ranked score is computed (Fig. 1B, right). A lower score means a worse SDOH and vice versa. We later evaluate model performance according to different SDoH subgroups split for each outcome (Fig. 6).

### Identifying correlations between features

Prior to applying ML models to the cleaned data, we conducted correlation analysis between all features/SDoH and all adverse outcomes. Features were aggregated across the entire pregnancy, using their minimum, maximum, median, and mean values, in a format consistent with the input structure of the Logistic Regression and XGBoost models described below. We used multiple logistic regression adjusted for age, race, and smoking status to fit each feature and outcome pair and extracted corresponding odds ratio (indicating level of positive/negative correlation) and p-value. We applied a Bonferroni significance threshold adjusted for multiple comparisons as the level of significance: 3.45×10⁻ ⁶ (0.05 divided by the total number of tests, i.e., 2072 features × 7 outcomes = 14,504).

### Predicting pregnancy outcomes with ML models

With respect to each label, we formulated one binary classification problem (0=Normal Outcome, 1=Adverse Outcome). The data within an episode was selected from the label, from the start of pregnancy to some gap days (defaulted to be 28 days, 4 weeks) before the onset of the label. The normal outcome had all data 4 weeks prior to the delivery. We expected differences in window length to have limited influence. In the case of tabular models, data were aggregated across the entire window, regardless of the length of time series. In LSTM models, although the length of the time series varied, the absolute timestamps were not provided. Given the existing variability in data density across patients, we expected the differences in window length to remain effectively invisible to the models.

To benchmark our created dataset with our extracted label, we trained popular machine learning models, Logistic Regression, XGBoost, and a customized neural network-based LSTM model on different forms of the dataset, depending on the specifics of the model:

Logistic Regression: We transformed the data to tabular representations, where rows are each pregnancy episode and columns are features. To account for the variable number of feature occurrences during pregnancy period for different episodes (ranges from 0 to any), we compress multiple feature occurrences within the entire pregnancy interval. Mean, min, max, median were computed across pregnancy, so that the number of columns was equal to 4 times the number of features. Discrete values are one-hot encoded. There were large amounts of missing values, and we filled missing values with mean-imputation.

XGBoost: We performed similar pre-processing steps as in Logistic regression, but here missing values were directly passed in as NaN instead of imputing since XGBoost can accept missing values.

Embedded LSTM: To retain the original granularity and fully capture the complexity of the time series, we implemented an embedded LSTM model inspired by Rohan et al. (2020) [3] and Choi et al. (2016) [4]. The feature embedding was trained based on the name string of the feature using sentence transformer [5], and projected to a lower dimensionality of R^3 using T-SNE. Similar to [3], the temporal information was embedded as an R^2 vector using sinusoidal embeddings. Finally, the value of the feature was concatenated. Each feature occurrence is then represented as an R^6 vector in the sequence.

### Training and Evaluation

Train-Val-Test splits and Cross Validation: We used a train-val-test split of 60-15-25 and 4-fold cross validation. For each outcome and model, 4 separate trainings were performed with each 25 percent of samples used as hold-out test sets. The number four is chosen to balance the tradeoff between maximizing validation robustness and maintaining sufficient positive label counts within each subgroup, as positive cases are scarce for certain adverse outcomes.. General Training: To address the high data imbalance, we used label weighting during training for all the models. The class weights were derived directly from the observed positive-to-negative class proportions within each training fold (Fig. 1c), ensuring that minority (positive) classes contributed proportionally more to the optimization objective. This approach mitigated the tendency of models to over-predict the majority class and improved sensitivity for rare outcomes, without requiring oversampling or synthetic data augmentation that might distort the temporal or clinical structure of the EHR. LSTM Training: We used 5e-4 learning rate, Adam Optimizer, an epoch size of 30, and exponential learning rate scheduling to train the LSTM models. These hyperparameters were selected based on empirical stability and performance in early experiments, and were found to provide consistent convergence without much overfitting. All training was performed on a Nvidia Tesla T4 GPU. Metrics used for evaluation: AUROC and F1 scores (macro-averaged) were used to evaluate the result of different models on different outcomes. Bias Mitigation: We explored bias-mitigation strategies across SDoH, age, and race subgroups. Ultimately, the only viable approach we integrated was to include these variables directly as model features. Because only a small subset of the cohort took the SDoH survey (21%), the positive label counts in each subgroup were even more scarce. We tested upsampling subgroups using methods such as SMOTE but yielded poor performance likely due to high dimensionality. Downsampling was also infeasible given the limited positive label numbers. We also considered calibration and temperature scaling but observed no noticeable improvements. These constraints represent an important limitation of our study, which we explicitly acknowledge. This is a limitation of our study which we acknowledge. Gap Analysis: We investigated the effect of different gap days (14 days, 28 days, 42 days, 56 days) on the model performance (Fig. 3d,g). The training pipeline remained the same.

### Feature Importance Analysis of model predictions

We conducted post-hoc feature importance analysis of the ML models to understand what features were valued in the prediction models, as well as comparing them to the important features as found by correlation. For the XGBoost model, the SHAP importance score of each feature was used, which provided additive and model-agnostic measures of each feature’s contribution to the prediction [44]. For LSTM, we replaced each feature with random noise when evaluating its importance and repeated this for every feature (leave-one-out strategy) to obtain an AUROC loss score for every feature when it was excluded in training. A more negative AUROC loss would indicate that the feature was more important since model performance was impacted heavily when this feature was excluded.

### Clinician rating of statistical and ML features

To evaluate the alignment between features identified through correlation analysis or machine learning and those used in clinical practice, four of the research team who are practicing physicians, were blinded to which features were statistically correlated versus important for model prediction. Prior to the rating process, we met with the physician team to discuss the expectations and requirements for this rating process, and additional instructions were provided to ensure consistency. Instructions and the full spreadsheet used for physician ratings on features can be found in the appendix F. After the rating sheets were collected, we additionally met with physicians to discuss the ratings and clinical implications of the extracted features.

## Conclusion

This study demonstrates the potential of the “All of Us” dataset for understanding and predicting adverse maternal health outcomes. We demonstrate that meaningful populational level patterns can be extracted, and high-performing machine learning models can be trained on this diverse, multi-site dataset, enabled by rigorous data preprocessing and problem formulation methods. Most important features identified through correlation analysis align with known clinical risk factors, reinforcing the robustness of the dataset and our preprocessing pipeline. Notably, we also discovered some unexpected associations that warrant further clinical investigation. While machine learning models successfully leverage many of these relevant features, they also rely on some with limited or no apparent correlation to outcomes. These findings, if validated, could inform new strategies for maternal care.

## Data Availability

Processed dataset and related code would be made available at time of publication.

## Acknowledgement

We gratefully acknowledge All of Us participants for their contributions, without whom this research would not have been possible. We also thank the National Institutes of Health’s All of Us Research Program for making available the participant data [and/or samples and/or cohort] examined in this study.

## Author Contributions

I.C. and M.A. conceptualized the study. A.Z., K.H, I.C., and M.A. supervised the study. H.Z. curated the dataset and conducted analysis. H.Z. prepared figures and tables. H.Z., I.C., and M.A. interpreted the results and wrote the manuscript. A.Z., S.F., B.G., and N.Y. interpreted the clinical implications of the results. All authors contributed to manuscript editing.

## Competing Interests Statement

The authors declare no competing interests.

## Funding Statement

This research received no funding support.

## Consent to Participate Declaration

All participants enrolled in the All of Us Research Program agreed to the “Consent to Join the All of Us Research Program” document [36].

## Ethics Approval Declaration

The protocols, informed consent, and other ethics-related materials in the All of Us Research Program are reviewed by the All of Us Institutional Review Board (IRB) [45].

## Notes

### Competing Interest Statement

The authors have declared no competing interest.

### Author Declarations

All of Us Institutional Review Board of National Institutes of Health gave ethical approval for this work

