## Supplementary Materials for "Machine Learning-Based Prediction of Maternal Morbidity across Heterogeneous Populations in the United States using Sequential Modeling of the *All of Us* Dataset"

### Appendices

#### Appendix A: Labels Mapping

```
grouped_items = {
    'gestational_diabetes': [
        'Gestational diabetes mellitus',
        'Gestational diabetes mellitus in childbirth',
        'Postpartum gestational diabetes mellitus',
        'Gestational diabetes mellitus, class A>1<',
        'Gestational diabetes mellitus, class A>2<',
        'Gestational diabetes mellitus complicating pregnancy'
    ],
    'cardiomyopathy': [
        'Cardiomyopathy',
        'Primary cardiomyopathy',
        'Hypertrophic cardiomyopathy',
        'Cardiomyopathy associated with another disorder',
        'Dilated cardiomyopathy secondary to peripartum heart disease',
        'Dilated cardiomyopathy',
        'Hypertrophic obstructive cardiomyopathy',
        'Takotsubo cardiomyopathy',
        'Postpartum cardiomyopathy',
        'Endomyocardial fibrosis',
        'Myocardial degeneration',
        'Restrictive cardiomyopathy',
        'Cardiomyopathy caused by drug',
        'Cardiomyopathy due to viral infection',
        'Dilated cardiomyopathy secondary to alcohol',
        'Restrictive cardiomyopathy with endomyocardial fibrosis',
        'Nutritional and metabolic cardiomyopathies',
        'Dilated cardiomyopathy secondary to drug'
    ],
    'pre_eclampsia': [
        'Pre-eclampsia',
        'Severe pre-eclampsia',
        'Severe pre-eclampsia - delivered',
        'HELLP syndrome',
        'Severe pre-eclampsia - not delivered',
        'Mild pre-eclampsia',
        'Severe pre-eclampsia with postnatal complication',
        'Postpartum pre-eclampsia',
        'Severe pre-eclampsia - delivered with postnatal complication',
        'Pre-eclampsia added to pre-existing hypertension'
    ],
    'preterm_labor': [
```

```
        'Premature labor',
        'Preterm labor without delivery',
        'Preterm labor in third trimester with preterm delivery in third trimester',
        'Preterm spontaneous labor with preterm delivery',
        'Preterm labor in second trimester with preterm delivery in second trimester',
        'Preterm labor with preterm delivery',
        'Preterm spontaneous labor with term delivery'
    ],
    'depression_anxiety': [
        'Anxiety disorder',
        'Posttraumatic stress disorder',
        'Generalized anxiety disorder',
        'Depressive disorder',
        'Major depression, single episode',
        'Recurrent major depression',
        'Moderate recurrent major depression',
        'Anxiety state',
        'Dysthymia',
        'Panic disorder without agoraphobia',
        'Severe recurrent major depression without psychotic features',
        'Recurrent major depressive episodes, moderate',
        'Recurrent major depression in partial remission',
        'Mild recurrent major depression',
        'Chronic post-traumatic stress disorder',
        'Bipolar affective disorder, current episode depression',
        'Acute stress disorder',
        'Moderate major depression, single episode',
        'Recurrent major depression in full remission',
        'Recurrent major depressive episodes, severe, with psychosis',
        'Recurrent major depressive episodes, mild',
        'Recurrent major depression in remission',
        'Anxiety',
        'Obsessive-compulsive disorder',
        'Social phobia',
        'Mild major depression, single episode',
        'Panic disorder with agoraphobia',
        'Schizoaffective disorder, depressive type',
        'Postpartum depression',
        'Mixed anxiety and depressive disorder',
        'Recurrent major depressive episodes',
        'Severe major depression, single episode, without psychotic features',
        'Single episode of major depression in full remission',
        'Organic anxiety disorder',
        'Panic disorder',
        'Major depression single episode, in partial remission',
        'Premenstrual dysphoric disorder',
        'Bipolar affective disorder, currently depressed, mild',
```

92 'Severe major depression, single episode, with psychotic features',
93 'Psychoactive substance-induced organic anxiety disorder',
94 'Recurrent depression',
95 'Severe recurrent major depression with psychotic features',
96 'Phobic disorder',
97 'Panic attack',
98 'Bipolar affective disorder, currently depressed, in full remission',
99 'Psychosis and severe depression co-occurrent and due to bipolar affective disorder',
100 'Claustrophobia',
101 'Dream anxiety disorder',
102 'Major depressive disorder',
103 'Fear of flying',
104 'Severe recurrent major depression',
105 'Reactive depression (situational)',
106 'Physical AND emotional exhaustion state',
107 'Phobia',
108 'Atypical depressive disorder',
109 'Generalized social phobia',
110 'Anxiety disorder of childhood OR adolescence',
111 'Separation anxiety disorder of childhood',
112 'Agoraphobia',
113 'Severe depression',
114 'Panic',
115 'Chronic depression',
116 'Seasonal affective disorder',
117 'Mild depression',
118 'Flying phobia',
119 'Agoraphobia without history of panic disorder without limited symptom attacks',
120 'Agoraphobia without history of panic disorder',
121 'Recurrent major depressive disorder in partial remission co-occurrent with anxiety',
122 'Acute post-trauma stress state',
123 'Chronic depressive personality disorder',
124 'Major depression in partial remission',
125 'Moderate major depression',
126 'Moderately severe recurrent major depression',
127 'Anxiety about body function or health',
128 'Depressive disorder in mother complicating pregnancy',
129 'Alcohol-induced anxiety disorder',
130 'Acute situational disturbance',
131 'Anxiety disorder due to a general medical condition',
132 'Simple phobia',
133 'Moderate depression',
134 'Severe major depression, single episode',
135 'Zoophobia',
136 'Moderately severe depression',
137 'Performance anxiety',
138 'Reactive depressive psychosis',

```
        'Minimal depression',
        'Moderate anxiety',
        'Major depression in full remission',
        'Chronic anxiety',
        'Posttraumatic stress disorder, delayed onset',
        'Sedative, hypnotic AND/OR anxiolytic-induced anxiety disorder',
        'Severe postnatal depression',
        'Anxiety attack',
        'Major depression in remission',
        'Moderately severe major depression',
        'Needle phobia',
        'Severe major depression without psychotic features',
        'Single major depressive episode, severe, with psychosis',
        'Severe major depression',
        'Separation anxiety',
        'Anxiety about treatment',
        'Mild major depression',
        'Primary dysthymia',
        'Mild anxiety',
        'Severe major depression with psychotic features',
        'Chronic stress disorder',
        'Maternity blues',
        'Bipolar affective disorder, currently depressed, moderate',
        'Recurrent major depressive disorder with postpartum onset'
    ],
    'miscarriage': [
        'Missed miscarriage',
        'Miscarriage without complication',
        'Recurrent miscarriage',
        'Incomplete inevitable miscarriage without complication',
        'Miscarriage',
        'Incomplete miscarriage with delayed or excessive hemorrhage',
        'Miscarriage with complication',
        'Incomplete miscarriage with complication',
        'Complete miscarriage',
        'Incomplete miscarriage',
        'Miscarriage complicated by genital-pelvic infection',
        'Miscarriage with sepsis',
        'Miscarriage with heavy bleeding',
        'Miscarriage in first trimester',
        'Inevitable miscarriage',
        'Urinary tract infection due to incomplete miscarriage',
        'Incomplete miscarriage with damage to pelvic organs or tissues',
        'Complete inevitable miscarriage without complication',
        'Vanishing twin syndrome',
        'Incomplete miscarriage with embolism',
        'Miscarriage complicated by shock',
```

```
        'Complete miscarriage with metabolic disorder',
        'Miscarriage complicated by embolism',
        'Complete miscarriage with genital tract or pelvic infection',
        'Chemical pregnancy',
        'Miscarriage complicated by damage to pelvic organs and/or tissues',
        'Incomplete miscarriage with renal failure',
        'Complete miscarriage with shock',
        'Complete miscarriage with renal failure',
        'Incomplete miscarriage with genital tract or pelvic infection',
        'Complete miscarriage with delayed or excessive hemorrhage',
        'Miscarriage with urinary tract infection',
        'Incomplete miscarriage with metabolic disorder',
        'Miscarriage in second trimester'
    ],
    'eclampsia': [
        'Eclampsia',
        'Eclampsia - delivered',
        'Eclampsia - delivered with postnatal complication',
        'Eclampsia in labor',
        'Eclampsia in pregnancy',
        'Eclampsia in puerperium'
    ],}
```

#### Appendix B: Survey Question Mappings

The SDoH survey of All of Us Research Program is compiled based on 14 various sources. To unify them, we defined 6 categories below and mapped all survey questions to each of the categories and answers to a continuous value from -3 to 3. The definitions are:

- **Companion:** The degree one can feel and obtain support from other people close in their lives
- **Discrimination:** The frequency of one encountering discriminative events
- **Efficacy:** The feeling and confidence of one's ability to control their own life and continuous wellbeing without being disturbed significantly
- **Healthcare:** The access to healthcare resources and the quality of healthcare service received
- **Neighborhood:** Whether one lives in a community that makes them feel safe and included, and also contains good infrastructures such as public transportation systems
- **Religion:** The degree to which one can feel connected to the higher power in the religion they believe

| Question | answer | Category | Higher Better? | Value |
|---|---|---|---|---|
| How often do you feel isolated from others? | Never | Companion | FALSE | -3 |
| | Rarely | Companion | FALSE | -2 |
| | PMI: Skip | Companion | FALSE | 0 |
| | Sometimes | Companion | FALSE | 0 |
| | Often | Companion | FALSE | 2 |
| How often do you feel lack companionship? | Never | Companion | FALSE | -3 |
| | Rarely | Companion | FALSE | -2 |
| | PMI: Skip | Companion | FALSE | 0 |
| | Sometimes | Companion | FALSE | 0 |
| | Often | Companion | FALSE | 2 |
| How often do you feel left out? | Never | Companion | FALSE | -3 |
| | Rarely | Companion | FALSE | -2 |
| | PMI: Skip | Companion | FALSE | 0 |
| | Sometimes | Companion | FALSE | 0 |
| | Often | Companion | FALSE | 2 |
| How often do you feel that people are around you but not with you? | Never | Companion | FALSE | -3 |
| | Rarely | Companion | FALSE | -2 |
| | PMI: Skip | Companion | FALSE | 0 |
| | Sometimes | Companion | FALSE | 0 |
| | Often | Companion | FALSE | 2 |
| | Never | Companion | FALSE | -3 |

| | | | | |
|---|---|---|---|---|
| How often do you feel that there is no one you can turn to? | Rarely | Companion | FALSE | -2 |
| | PMI: Skip | Companion | FALSE | 0 |
| | Sometimes | Companion | FALSE | 0 |
| | Often | Companion | FALSE | 2 |
| How often do you feel that you are an outgoing person? | Never | Companion | TRUE | -3 |
| | Rarely | Companion | TRUE | -2 |
| | PMI: Skip | Companion | TRUE | 0 |
| | Sometimes | Companion | TRUE | 0 |
| | Often | Companion | TRUE | 2 |
| How often do you feel that you are unhappy being so withdrawn? | Never | Companion | FALSE | -3 |
| | Rarely | Companion | FALSE | -2 |
| | PMI: Skip | Companion | FALSE | 0 |
| | Sometimes | Companion | FALSE | 0 |
| | Often | Companion | FALSE | 2 |
| How often do you fell that you can find companionship when you want it? | Never | Companion | TRUE | -3 |
| | Rarely | Companion | TRUE | -2 |
| | PMI: Skip | Companion | TRUE | 0 |
| | Sometimes | Companion | TRUE | 0 |
| | Often | Companion | TRUE | 2 |
| How often do you have someone to have a good time with? | None of the time | Companion | TRUE | -3 |
| | A little of the time | Companion | TRUE | -2 |
| | PMI: Skip | Companion | TRUE | 0 |
| | Some of the time | Companion | TRUE | 0 |
| | Most of the time | Companion | TRUE | 2 |
| | All of the time | Companion | TRUE | 3 |
| How often do you have someone to help you if you were confined to bed? | None of the time | Companion | TRUE | -3 |
| | A little of the time | Companion | TRUE | -2 |
| | PMI: Skip | Companion | TRUE | 0 |
| | Some of the time | Companion | TRUE | 0 |
| | Most of the time | Companion | TRUE | 2 |
| | All of the time | Companion | TRUE | 3 |
| How often do you have someone to help you with daily chores if you were sick? | None of the time | Companion | TRUE | -3 |
| | A little of the time | Companion | TRUE | -2 |
| | PMI: Skip | Companion | TRUE | 0 |

| | | | | |
|---|---|---|---|---|
| | Some of the time | Companion | TRUE | 0 |
| | Most of the time | Companion | TRUE | 2 |
| | All of the time | Companion | TRUE | 3 |
| How often do you have someone to love and make you feel wanted? | None of the time | Companion | TRUE | -3 |
| | A little of the time | Companion | TRUE | -2 |
| | PMI: Skip | Companion | TRUE | 0 |
| | Some of the time | Companion | TRUE | 0 |
| | Most of the time | Companion | TRUE | 2 |
| | All of the time | Companion | TRUE | 3 |
| How often do you have someone to prepare your meals if you were unable to do it yourself? | None of the time | Companion | TRUE | -3 |
| | A little of the time | Companion | TRUE | -2 |
| | PMI: Skip | Companion | TRUE | 0 |
| | Some of the time | Companion | TRUE | 0 |
| | Most of the time | Companion | TRUE | 2 |
| | All of the time | Companion | TRUE | 3 |
| How often do you have someone to take you to the doctor if you need it? | None of the time | Companion | TRUE | -3 |
| | A little of the time | Companion | TRUE | -2 |
| | PMI: Skip | Companion | TRUE | 0 |
| | Some of the time | Companion | TRUE | 0 |
| | Most of the time | Companion | TRUE | 2 |
| | All of the time | Companion | TRUE | 3 |
| How often do you have someone to turn to for suggestions about how to deal with a personal problem? | None of the time | Companion | TRUE | -3 |
| | A little of the time | Companion | TRUE | -2 |
| | PMI: Skip | Companion | TRUE | 0 |
| | Some of the time | Companion | TRUE | 0 |
| | Most of the time | Companion | TRUE | 2 |
| | All of the time | Companion | TRUE | 3 |
| How often do you have someone who understands your problems? | None of the time | Companion | TRUE | -3 |
| | A little of the time | Companion | TRUE | -2 |
| | PMI: Skip | Companion | TRUE | 0 |
| | Some of the time | Companion | TRUE | 0 |
| | Most of the time | Companion | TRUE | 2 |
| | All of the time | Companion | TRUE | 3 |
| | Never | Discrimination | FALSE | -3 |

| | | | | |
|---|---|---|---|---|
| In your day-to-day life, how often are you called names or insulted? | Less than once a year | Discrimination | FALSE | -2 |
| | A few times a year | Discrimination | FALSE | -1 |
| | PMI: Skip | Discrimination | FALSE | 0 |
| | A few times a month | Discrimination | FALSE | 1 |
| | At least once a week | Discrimination | FALSE | 2 |
| | Almost everyday | Discrimination | FALSE | 3 |
| In your day-to-day life, how often are you threatened or harassed? | Never | Discrimination | FALSE | -3 |
| | Less than once a year | Discrimination | FALSE | -2 |
| | A few times a year | Discrimination | FALSE | -1 |
| | PMI: Skip | Discrimination | FALSE | 0 |
| | A few times a month | Discrimination | FALSE | 1 |
| | At least once a week | Discrimination | FALSE | 2 |
| | Almost everyday | Discrimination | FALSE | 3 |
| In your day-to-day life, how often are you treated with less courtesy than other people? | Never | Discrimination | FALSE | -3 |
| | Less than once a year | Discrimination | FALSE | -2 |
| | A few times a year | Discrimination | FALSE | -1 |
| | PMI: Skip | Discrimination | FALSE | 0 |
| | A few times a month | Discrimination | FALSE | 1 |
| | At least once a week | Discrimination | FALSE | 2 |
| | Almost everyday | Discrimination | FALSE | 3 |
| In your day-to-day life, how often are you treated with less respect than other people? | Never | Discrimination | FALSE | -3 |
| | Less than once a year | Discrimination | FALSE | -2 |
| | A few times a year | Discrimination | FALSE | -1 |
| | PMI: Skip | Discrimination | FALSE | 0 |
| | A few times a month | Discrimination | FALSE | 1 |
| | At least once a week | Discrimination | FALSE | 2 |
| | Almost everyday | Discrimination | FALSE | 3 |
| In your day-to-day life, how often do people act as if they are afraid of you? | Never | Discrimination | FALSE | -3 |
| | Less than once a year | Discrimination | FALSE | -2 |
| | A few times a year | Discrimination | FALSE | -1 |
| | PMI: Skip | Discrimination | FALSE | 0 |
| | A few times a month | Discrimination | FALSE | 1 |
| | At least once a week | Discrimination | FALSE | 2 |

| | | | | |
|---|---|---|---|---|
| | Almost everyday | Discrimination | FALSE | 3 |
| In your day-to-day life, how often do people act as if they think you are dishonest? | Never | Discrimination | FALSE | -3 |
| | Less than once a year | Discrimination | FALSE | -2 |
| | A few times a year | Discrimination | FALSE | -1 |
| | PMI: Skip | Discrimination | FALSE | 0 |
| | A few times a month | Discrimination | FALSE | 1 |
| | At least once a week | Discrimination | FALSE | 2 |
| | Almost everyday | Discrimination | FALSE | 3 |
| In your day-to-day life, how often do people act as if they think you are not smart? | Never | Discrimination | FALSE | -3 |
| | Less than once a year | Discrimination | FALSE | -2 |
| | A few times a year | Discrimination | FALSE | -1 |
| | PMI: Skip | Discrimination | FALSE | 0 |
| | A few times a month | Discrimination | FALSE | 1 |
| | At least once a week | Discrimination | FALSE | 2 |
| | Almost everyday | Discrimination | FALSE | 3 |
| In your day-to-day life, how often do people act as if they're better than you are? | Never | Discrimination | FALSE | -3 |
| | Less than once a year | Discrimination | FALSE | -2 |
| | A few times a year | Discrimination | FALSE | -1 |
| | PMI: Skip | Discrimination | FALSE | 0 |
| | A few times a month | Discrimination | FALSE | 1 |
| | At least once a week | Discrimination | FALSE | 2 |
| | Almost everyday | Discrimination | FALSE | 3 |
| In your day-to-day life, how often do you receive poorer service than other people at restaurants or stores? | Never | Discrimination | FALSE | -3 |
| | Less than once a year | Discrimination | FALSE | -2 |
| | A few times a year | Discrimination | FALSE | -1 |
| | PMI: Skip | Discrimination | FALSE | 0 |
| | A few times a month | Discrimination | FALSE | 1 |
| | At least once a week | Discrimination | FALSE | 2 |
| | Almost everyday | Discrimination | FALSE | 3 |
| How often do you feel deep inner peace or harmony? | Never or almost never | Efficacy | TRUE | -3 |
| | Once in a while | Efficacy | TRUE | -1 |
| | PMI: Skip | Efficacy | TRUE | 0 |
| | Some days | Efficacy | TRUE | 1 |

| | | | | |
|---|---|---|---|---|
| | Many times a day | Efficacy | TRUE | 2 |
| | Most days | Efficacy | TRUE | 2 |
| | Every day | Efficacy | TRUE | 3 |
| In the last month, how often have you been able to control irritations in your life? | Almost Never | Efficacy | TRUE | -3 |
| | Never | Efficacy | TRUE | -3 |
| | PMI: Skip | Efficacy | TRUE | 0 |
| | Sometimes | Efficacy | TRUE | 0 |
| | Fairly Often | Efficacy | TRUE | 2 |
| | Very Often | Efficacy | TRUE | 3 |
| In the last month, how often have you been angered because of things that were outside of your control? | Almost Never | Efficacy | FALSE | -3 |
| | Never | Efficacy | FALSE | -3 |
| | PMI: Skip | Efficacy | FALSE | 0 |
| | Sometimes | Efficacy | FALSE | 0 |
| | Fairly Often | Efficacy | FALSE | 2 |
| | Very Often | Efficacy | FALSE | 3 |
| In the last month, how often have you been upset because of something that happened unexpectedly? | Almost Never | Efficacy | FALSE | -3 |
| | Never | Efficacy | FALSE | -3 |
| | PMI: Skip | Efficacy | FALSE | 0 |
| | Sometimes | Efficacy | FALSE | 0 |
| | Fairly Often | Efficacy | FALSE | 2 |
| | Very Often | Efficacy | FALSE | 3 |
| In the last month, how often have you felt confident about your ability to handle your personal problems? | Almost Never | Efficacy | FALSE | -3 |
| | Never | Efficacy | FALSE | -3 |
| | PMI: Skip | Efficacy | FALSE | 0 |
| | Sometimes | Efficacy | FALSE | 0 |
| | Fairly Often | Efficacy | FALSE | 2 |
| | Very Often | Efficacy | FALSE | 3 |
| In the last month, how often have you felt difficulties were piling up so high that you could not overcome them? | Almost Never | Efficacy | FALSE | -3 |
| | Never | Efficacy | FALSE | -3 |
| | PMI: Skip | Efficacy | FALSE | 0 |
| | Sometimes | Efficacy | FALSE | 0 |
| | Fairly Often | Efficacy | FALSE | 2 |
| | Very Often | Efficacy | FALSE | 3 |
| | Almost Never | Efficacy | FALSE | -3 |

| | | | | |
|---|---|---|---|---|
| In the last month, how often have you felt nervous and "stressed"? | Never | Efficacy | FALSE | -3 |
| | PMI: Skip | Efficacy | FALSE | 0 |
| | Sometimes | Efficacy | FALSE | 0 |
| | Fairly Often | Efficacy | FALSE | 2 |
| | Very Often | Efficacy | FALSE | 3 |
| In the last month, how often have you felt that things were going your way? | Almost Never | Efficacy | FALSE | -3 |
| | Never | Efficacy | FALSE | -3 |
| | PMI: Skip | Efficacy | FALSE | 0 |
| | Sometimes | Efficacy | FALSE | 0 |
| | Fairly Often | Efficacy | FALSE | 2 |
| | Very Often | Efficacy | FALSE | 3 |
| In the last month, how often have you felt that you were on top of things? | Almost Never | Efficacy | FALSE | -3 |
| | Never | Efficacy | FALSE | -3 |
| | PMI: Skip | Efficacy | FALSE | 0 |
| | Sometimes | Efficacy | FALSE | 0 |
| | Fairly Often | Efficacy | FALSE | 2 |
| | Very Often | Efficacy | FALSE | 3 |
| In the last month, how often have you felt that you were unable to control the important things in your life? | Almost Never | Efficacy | FALSE | -3 |
| | Never | Efficacy | FALSE | -3 |
| | PMI: Skip | Efficacy | FALSE | 0 |
| | Sometimes | Efficacy | FALSE | 0 |
| | Fairly Often | Efficacy | FALSE | 2 |
| | Very Often | Efficacy | FALSE | 3 |
| In the last month, how often have you found that you could not cope with all the things that you had to do? | Almost Never | Efficacy | FALSE | -3 |
| | Never | Efficacy | FALSE | -3 |
| | PMI: Skip | Efficacy | FALSE | 0 |
| | Sometimes | Efficacy | FALSE | 0 |
| | Fairly Often | Efficacy | FALSE | 2 |
| | Very Often | Efficacy | FALSE | 3 |
| Within the past 12 months, were you worried whether the food you had bought just didn't last and you didn't have money to get more? | Never true | Efficacy | FALSE | -3 |
| | PMI: Skip | Efficacy | FALSE | 0 |
| | Sometimes true | Efficacy | FALSE | 1 |
| | Often true | Efficacy | FALSE | 2 |
| | Never true | Efficacy | FALSE | -3 |

| | | | | |
|---|---|---|---|---|
| Within the past 12 months, were you worried whether your food would run out before you got money to buy more? | PMI: Skip | Efficacy | FALSE | 0 |
| | Sometimes true | Efficacy | FALSE | 1 |
| | Often true | Efficacy | FALSE | 2 |
| How often are you treated with less courtesy than other people when you go to a doctor's office or other health care provider? | Never | Healthcare | FALSE | -3 |
| | Rarely | Healthcare | FALSE | -2 |
| | PMI: Skip | Healthcare | FALSE | 0 |
| | Sometimes | Healthcare | FALSE | 0 |
| | Most of the time | Healthcare | FALSE | 2 |
| How often are you treated with less respect than other people when you go to a doctor's office or other health care provider? | Never | Healthcare | FALSE | -3 |
| | Rarely | Healthcare | FALSE | -2 |
| | PMI: Skip | Healthcare | FALSE | 0 |
| | Sometimes | Healthcare | FALSE | 0 |
| | Most of the time | Healthcare | FALSE | 2 |
| How often do you feel like a doctor or nurse is not listening to what you were saying. when you go to a doctor's office or other health care provider? | Never | Healthcare | FALSE | -3 |
| | Rarely | Healthcare | FALSE | -2 |
| | PMI: Skip | Healthcare | FALSE | 0 |
| | Sometimes | Healthcare | FALSE | 0 |
| | Most of the time | Healthcare | FALSE | 2 |
| How often do you receive poorer service than others when you go to a doctor's office or other health care provider? | Never | Healthcare | FALSE | -3 |
| | Rarely | Healthcare | FALSE | -2 |
| | PMI: Skip | Healthcare | FALSE | 0 |
| | Sometimes | Healthcare | FALSE | 0 |
| | Most of the time | Healthcare | FALSE | 2 |
| How often does a doctor or nurse act as if he or she is afraid of you when you go to a doctor's office or other health care provider? | Never | Healthcare | FALSE | -3 |
| | Rarely | Healthcare | FALSE | -2 |
| | PMI: Skip | Healthcare | FALSE | 0 |
| | Sometimes | Healthcare | FALSE | 0 |
| | Most of the time | Healthcare | FALSE | 2 |
| How often does a doctor or nurse act as if he or she is better than you when you go to a doctor's office or other health care provider? | Never | Healthcare | FALSE | -3 |
| | Rarely | Healthcare | FALSE | -2 |
| | PMI: Skip | Healthcare | FALSE | 0 |
| | Sometimes | Healthcare | FALSE | 0 |
| | Most of the time | Healthcare | FALSE | 2 |
| | Never | Healthcare | FALSE | -3 |

| | | | | |
|---|---|---|---|---|
| How often does a doctor or nurse act as if he or she thinks you are not smart when you go to a doctor's office or other health care provider? | Rarely | Healthcare | FALSE | -2 |
| | PMI: Skip | Healthcare | FALSE | 0 |
| | Sometimes | Healthcare | FALSE | 0 |
| | Most of the time | Healthcare | FALSE | 2 |
| How much you agree or disagree that in your neighborhood people watch out for each other? | Strongly disagree | Neighborhood | TRUE | -3 |
| | Disagree | Neighborhood | TRUE | -2 |
| | PMI: Skip | Neighborhood | TRUE | 0 |
| | Agree | Neighborhood | TRUE | 2 |
| | Strongly agree | Neighborhood | TRUE | 3 |
| How much you agree or disagree that people around here are willing to help their neighbor? | Strongly disagree | Neighborhood | TRUE | -3 |
| | Disagree | Neighborhood | TRUE | -2 |
| | Neutral (neither agree nor disagree) | Neighborhood | TRUE | 0 |
| | PMI: Skip | Neighborhood | TRUE | 0 |
| | Agree | Neighborhood | TRUE | 2 |
| | Strongly agree | Neighborhood | TRUE | 3 |
| How much you agree or disagree that people in your neighborhood can be trusted? | Strongly disagree | Neighborhood | TRUE | -3 |
| | Disagree | Neighborhood | TRUE | -2 |
| | Neutral (neither agree nor disagree) | Neighborhood | TRUE | 0 |
| | PMI: Skip | Neighborhood | TRUE | 0 |
| | Agree | Neighborhood | TRUE | 2 |
| | Strongly agree | Neighborhood | TRUE | 3 |
| How much you agree or disagree that people in your neighborhood generally get along with each other? | Strongly disagree | Neighborhood | TRUE | -3 |
| | Disagree | Neighborhood | TRUE | -2 |
| | Neutral (neither agree nor disagree) | Neighborhood | TRUE | 0 |
| | PMI: Skip | Neighborhood | TRUE | 0 |
| | Agree | Neighborhood | TRUE | 2 |
| | Strongly agree | Neighborhood | TRUE | 3 |
| How much you agree or disagree that people in your neighborhood share the same values? | Strongly disagree | Neighborhood | TRUE | -3 |
| | Disagree | Neighborhood | TRUE | -2 |
| | Neutral (neither agree nor disagree) | Neighborhood | TRUE | 0 |
| | PMI: Skip | Neighborhood | TRUE | 0 |
| | Agree | Neighborhood | TRUE | 2 |
| | Strongly agree | Neighborhood | TRUE | 3 |
| | Strongly disagree | Neighborhood | TRUE | -3 |

| | | | | |
|---|---|---|---|---|
| How much you agree or disagree that people in your neighborhood take good care of their houses and apartments? | Disagree | Neighborhood | TRUE | -2 |
| | PMI: Skip | Neighborhood | TRUE | 0 |
| | Agree | Neighborhood | TRUE | 2 |
| | Strongly agree | Neighborhood | TRUE | 3 |
| How much you agree or disagree that there are lot of abandoned buildings in your neighborhood? | Strongly disagree | Neighborhood | FALSE | -3 |
| | Disagree | Neighborhood | FALSE | -2 |
| | PMI: Skip | Neighborhood | FALSE | 0 |
| | Agree | Neighborhood | FALSE | 2 |
| | Strongly agree | Neighborhood | FALSE | 3 |
| How much you agree or disagree that there are too many people hanging around on the streets near your home? | Strongly disagree | Neighborhood | FALSE | -3 |
| | Disagree | Neighborhood | FALSE | -2 |
| | PMI: Skip | Neighborhood | FALSE | 0 |
| | Agree | Neighborhood | FALSE | 2 |
| | Strongly agree | Neighborhood | FALSE | 3 |
| How much you agree or disagree that there is a lot of crime in your neighborhood? | Strongly disagree | Neighborhood | FALSE | -3 |
| | Disagree | Neighborhood | FALSE | -2 |
| | PMI: Skip | Neighborhood | FALSE | 0 |
| | Agree | Neighborhood | FALSE | 2 |
| | Strongly agree | Neighborhood | FALSE | 3 |
| How much you agree or disagree that there is a lot of graffiti in your neighborhood? | Strongly disagree | Neighborhood | FALSE | -3 |
| | Disagree | Neighborhood | FALSE | -2 |
| | PMI: Skip | Neighborhood | FALSE | 0 |
| | Agree | Neighborhood | FALSE | 2 |
| | Strongly agree | Neighborhood | FALSE | 3 |
| How much you agree or disagree that there is too much alcohol use in your neighborhood? | Strongly disagree | Neighborhood | FALSE | -3 |
| | Disagree | Neighborhood | FALSE | -2 |
| | PMI: Skip | Neighborhood | FALSE | 0 |
| | Agree | Neighborhood | FALSE | 2 |
| | Strongly agree | Neighborhood | FALSE | 3 |
| How much you agree or disagree that there is too much drug use in your neighborhood? | Strongly disagree | Neighborhood | FALSE | -3 |
| | Disagree | Neighborhood | FALSE | -2 |
| | PMI: Skip | Neighborhood | FALSE | 0 |
| | Agree | Neighborhood | FALSE | 2 |
| | Strongly agree | Neighborhood | FALSE | 3 |

| | | | | |
|---|---|---|---|---|
| How much you agree or disagree that vandalism is common in your neighborhood? | Strongly disagree | Neighborhood | FALSE | -3 |
| | Disagree | Neighborhood | FALSE | -2 |
| | PMI: Skip | Neighborhood | FALSE | 0 |
| | Agree | Neighborhood | FALSE | 2 |
| | Strongly agree | Neighborhood | FALSE | 3 |
| How much you agree or disagree that you are always having trouble with your neighbors? | Strongly disagree | Neighborhood | FALSE | -3 |
| | Disagree | Neighborhood | FALSE | -2 |
| | PMI: Skip | Neighborhood | FALSE | 0 |
| | Agree | Neighborhood | FALSE | 2 |
| | Strongly agree | Neighborhood | FALSE | 3 |
| How much you agree or disagree that your neighborhood is clean? | Strongly disagree | Neighborhood | TRUE | -3 |
| | Disagree | Neighborhood | TRUE | -2 |
| | PMI: Skip | Neighborhood | TRUE | 0 |
| | Agree | Neighborhood | TRUE | 2 |
| | Strongly agree | Neighborhood | TRUE | 3 |
| How much you agree or disagree that your neighborhood is noisy? | Strongly disagree | Neighborhood | FALSE | -3 |
| | Disagree | Neighborhood | FALSE | -2 |
| | PMI: Skip | Neighborhood | FALSE | 0 |
| | Agree | Neighborhood | FALSE | 2 |
| | Strongly agree | Neighborhood | FALSE | 3 |
| How much you agree or disagree that your neighborhood is safe? | Strongly disagree | Neighborhood | TRUE | -3 |
| | Disagree | Neighborhood | TRUE | -2 |
| | PMI: Skip | Neighborhood | TRUE | 0 |
| | Agree | Neighborhood | TRUE | 2 |
| | Strongly agree | Neighborhood | TRUE | 3 |
| It is within a 10-15 minute walk to a transit stop (such as bus, train, trolley, or tram) from my home. Would you say that you... | Strongly disagree | Neighborhood | TRUE | -3 |
| | Somewhat disagree | Neighborhood | TRUE | -1 |
| | PMI: Dont Know | Neighborhood | TRUE | 0 |
| | PMI: Skip | Neighborhood | TRUE | 0 |
| | Somewhat agree | Neighborhood | TRUE | 1 |
| | Strongly agree | Neighborhood | TRUE | 3 |
| Many shops, stores, markets or other places to buy things I need are within easy walking distance of my home. Would you say that you... | Strongly disagree | Neighborhood | TRUE | -3 |
| | Somewhat disagree | Neighborhood | TRUE | -1 |
| | PMI: Skip | Neighborhood | TRUE | 0 |

| | | | | |
|---|---|---|---|---|
| | PMI: Dont Know | Neighborhood | TRUE | 0 |
| | Somewhat agree | Neighborhood | TRUE | 1 |
| | Strongly agree | Neighborhood | TRUE | 3 |
| My neighborhood has several free or low-cost recreation facilities, such as parks, walking trails, bike paths, recreation centers, playgrounds, public swimming pools, etc. Would you say that you... | Strongly disagree | Neighborhood | TRUE | -3 |
| | Somewhat disagree | Neighborhood | TRUE | -1 |
| | PMI: Dont Know | Neighborhood | TRUE | 0 |
| | PMI: Skip | Neighborhood | TRUE | 0 |
| | Somewhat agree | Neighborhood | TRUE | 1 |
| | Strongly agree | Neighborhood | TRUE | 3 |
| The crime rate in my neighborhood makes it unsafe to go on walks at night. Would you say that you... | Strongly disagree | Neighborhood | FALSE | -3 |
| | Somewhat disagree | Neighborhood | FALSE | -1 |
| | PMI: Dont Know | Neighborhood | FALSE | 0 |
| | PMI: Skip | Neighborhood | FALSE | 0 |
| | Somewhat agree | Neighborhood | FALSE | 1 |
| | Strongly agree | Neighborhood | FALSE | 3 |
| The crime rate in my neighborhood makes it unsafe to go on walks during the day. Would you say that you... | Strongly disagree | Neighborhood | FALSE | -3 |
| | Somewhat disagree | Neighborhood | FALSE | -1 |
| | PMI: Skip | Neighborhood | FALSE | 0 |
| | PMI: Dont Know | Neighborhood | FALSE | 0 |
| | Somewhat agree | Neighborhood | FALSE | 1 |
| | Strongly agree | Neighborhood | FALSE | 3 |
| There are facilities to bicycle in or near my neighborhood, such as special lanes, separate paths or trails, or shared use paths for cycles and pedestrians. Would you say that you... | Strongly disagree | Neighborhood | TRUE | -3 |
| | Somewhat disagree | Neighborhood | TRUE | -1 |
| | Does not apply to my neighborhood | Neighborhood | TRUE | 0 |
| | PMI: Skip | Neighborhood | TRUE | 0 |
| | PMI: Dont Know | Neighborhood | TRUE | 0 |
| | Somewhat agree | Neighborhood | TRUE | 1 |
| | Strongly agree | Neighborhood | TRUE | 3 |
| There are sidewalks on most of the streets in my neighborhood. Would you say that you... | Strongly disagree | Neighborhood | TRUE | -3 |
| | Somewhat disagree | Neighborhood | TRUE | -1 |
| | Does not apply to my neighborhood | Neighborhood | TRUE | 0 |
| | PMI: Dont Know | Neighborhood | TRUE | 0 |
| | PMI: Skip | Neighborhood | TRUE | 0 |

| | | | | |
|---|---|---|---|---|
| | Somewhat agree | Neighborhood | TRUE | 1 |
| | Strongly agree | Neighborhood | TRUE | 3 |
| How often do you desire to be closer to or in union with God (or a higher power)? | Never or almost never | Religion | FALSE | -3 |
| | Once in a while | Religion | FALSE | -1 |
| | I do not believe in God (or a higher power) | Religion | FALSE | 0 |
| | PMI: Skip | Religion | FALSE | 0 |
| | Some days | Religion | FALSE | 1 |
| | Many times a day | Religion | FALSE | 2 |
| | Most days | Religion | FALSE | 2 |
| | Every day | Religion | FALSE | 3 |
| How often do you feel God's (or a higher power's) love for you, directly or through others? | Never or almost never | Religion | TRUE | -3 |
| | Once in a while | Religion | TRUE | -1 |
| | I do not believe in God (or a higher power) | Religion | TRUE | 0 |
| | PMI: Skip | Religion | TRUE | 0 |
| | Some days | Religion | TRUE | 1 |
| | Many times a day | Religion | TRUE | 2 |
| | Most days | Religion | TRUE | 2 |
| | Every day | Religion | TRUE | 3 |
| How often do you feel God's (or a higher power's) presence? | Never or almost never | Religion | TRUE | -3 |
| | Once in a while | Religion | TRUE | -1 |
| | I do not believe in God (or a higher power) | Religion | TRUE | 0 |
| | PMI: Skip | Religion | TRUE | 0 |
| | Some days | Religion | TRUE | 1 |
| | Many times a day | Religion | TRUE | 2 |
| | Most days | Religion | TRUE | 2 |
| | Every day | Religion | TRUE | 3 |
| How often do you feel that you are spiritually touched by the beauty of creation? | Never or almost never | Religion | TRUE | -3 |
| | Once in a while | Religion | TRUE | -1 |
| | PMI: Skip | Religion | TRUE | 0 |
| | Some days | Religion | TRUE | 1 |
| | Many times a day | Religion | TRUE | 2 |
| | Most days | Religion | TRUE | 2 |
| | Every day | Religion | TRUE | 3 |

| | | | | |
|---|---|---|---|---|
| How often do you find strength and comfort in your religion? | Never or almost never | Religion | TRUE | -3 |
| | Once in a while | Religion | TRUE | -1 |
| | I am not religious | Religion | TRUE | 0 |
| | PMI: Skip | Religion | TRUE | 0 |
| | Some days | Religion | TRUE | 1 |
| | Many times a day | Religion | TRUE | 2 |
| | Most days | Religion | TRUE | 2 |
| | Every day | Religion | TRUE | 3 |
| How often do you go to religious meetings or services? | Never or almost never | Religion | TRUE | -3 |
| | Less than once per month | Religion | TRUE | -1 |
| | I am not religious | Religion | TRUE | 0 |
| | PMI: Skip | Religion | TRUE | 0 |
| | 1 to 3 times per month | Religion | TRUE | 1 |
| | Once a week | Religion | TRUE | 1 |
| | More than once a week | Religion | TRUE | 2 |

### Appendix C: Dominant Unit

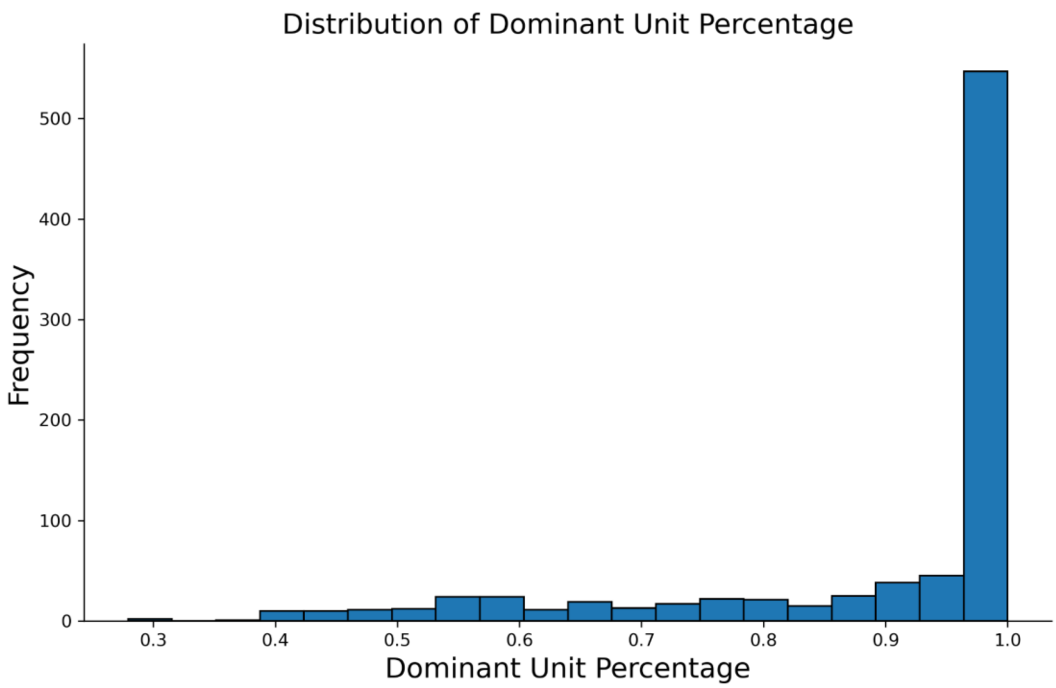

**Supplementary Figure 1** A histogram showing, for 1,000 features, the percentage of rows belonging to each feature's dominant unit.

| Potentially Collapsable Features, from top 1000 features, sorted A-Z | | | | | |
|---|---|---|---|---|---|
| **Glucose** | **Leukocytes** | **Hemoglobin** | **Hepatitis** | **HIV** | **Erythrocytes** |
| Abnormal glucose level', | Band form neutrophils/100 leukocytes in Blood by Manual count', | Glucose mean value [Mass/volume] in Blood Estimated from glycated hemoglobin', | Chronic hepatitis C', | HIV 1 Ab [Presence] in Serum or Plasma by Immunoassay', | Erythrocytes [#/area] in Urine sediment by Automated count', |
| Blood glucose abnormal', | Band form neutrophils/100 leukocytes in Blood', | Hemoglobin [Entitic mass] in Reticulocytes by Automated count', | Hepatitis A virus IgM Ab [Presence] in Serum or Plasma by Immunoassay'] | HIV 1 Ab [Presence] in Serum', | Erythrocytes [#/area] in Urine sediment by Microscopy high power field', |
| Fasting glucose [Mass/volume] in Serum or Plasma', | Basophils/100 leukocytes in Blood by Automated count', | Hemoglobin [Mass/volume] in Blood', | Hepatitis B virus core Ab [Presence] in Serum or Plasma by Immunoassay', | HIV 1 Ab [Units/volume] in Serum or Plasma by Immunoassay', | Erythrocytes [#/volume] in Blood by Automated count', |
| Glucose [Mass/volume] in Blood', | Basophils/100 leukocytes in Blood by Manual count', | Hemoglobin [Mass/volume] in Capillary blood', | Hepatitis B virus surface Ab [Presence] in Serum by Immunoassay', | HIV 1 p24 Ag [Presence] in Serum or Plasma by Immunoassay', | Erythrocytes [#/volume] in Blood', |
| Glucose [Mass/volume] in Capillary blood by Glucometer', | Basophils/100 leukocytes in Blood', | Hemoglobin [Mass/volume] in Urine by Automated test strip', | Hepatitis B virus surface Ab [Presence] in Serum', | HIV 1+2 Ab [Presence] in Serum or Plasma by Immunoassay', | Erythrocytes [#/volume] in Urine by Automated test strip', |
| Glucose [Mass/volume] in Serum or Plasma --1 hour post 100 g glucose PO', | Basophils/100 leukocytes in Body fluid by Manual count', | Hemoglobin [Mass/volume] in Venous blood', | Hepatitis B virus surface Ab [Units/volume] in Serum by Radioimmunoassay (RIA)', | HIV 1+2 Ab [Presence] in Serum, Plasma or Blood by Rapid immunoassay', | Erythrocytes [#/volume] in Urine by Test strip', |
| Glucose [Mass/volume] in Serum or Plasma --1 hour post 50 g glucose PO', | Eosinophils/100 leukocytes in Blood by Automated count', | Hemoglobin [Presence] in Urine by Automated test strip', | Hepatitis B virus surface Ab [Units/volume] in Serum or Plasma by Immunoassay', | HIV 1+2 Ab [Presence] in Serum', | Erythrocytes [#/volume] in Urine sediment by Microscopy high power field', |
| Glucose [Mass/volume] in Serum or Plasma --1 hour post 75 g glucose PO', | Eosinophils/100 leukocytes in Blood by Manual count', | Hemoglobin [Presence] in Urine by Test strip', | Hepatitis B virus surface Ab [Units/volume] in Serum', | HIV 1+2 Ab [Presence] in Specimen by Immunoassay', | Erythrocytes [#/volume] in Urine', |
| Glucose [Mass/volume] in Serum or Plasma --1 hour post dose glucose', | Eosinophils/100 leukocytes in Blood', | Hemoglobin [Presence] in Urine', | Hepatitis B virus surface Ag [Presence] in Serum from Donor by Immunoassay', | HIV 1+2 Ab [Presence] in Specimen by Rapid immunoassay', | Erythrocytes [Presence] in Urine sediment by Light microscopy'] |
| Glucose [Mass/volume] in Serum or Plasma --1 hour post meal', | Immature granulocytes/100 leukocytes in Blood by Automated count', | Hemoglobin A/Hemoglobin.total in Blood by Electrophoresis', | Hepatitis B virus surface Ag [Presence] in Serum or Plasma by Immunoassay', | HIV 1+2 Ab and HIV1 p24 Ag [Identifier] in Serum, Plasma or Blood by Rapid immunoassay', | Erythrocytes [Presence] in Urine', |
| Glucose [Mass/volume] in Serum or Plasma --1 hour post XXX challenge', | Immature granulocytes/100 leukocytes in Blood', | Hemoglobin A/Hemoglobin.total in Blood by HPLC', | Hepatitis B virus surface Ag [Presence] in Serum or Plasma by Neutralization test', | HIV 1+2 Ab+HIV1 p24 Ag [Presence] in Serum or Plasma by Immunoassay', | Leukocytes [#/volume] corrected for nucleated erythrocytes in Blood by Automated count', |
| Glucose [Mass/volume] in Serum or Plasma --2 hours post 100 g glucose PO', | Leukocytes [#/area] in Urine sediment by Automated count', | Hemoglobin A/Hemoglobin.total in Blood', | Hepatitis B virus surface Ag [Presence] in Serum', | HIV 1+O+2 Ab [Presence] in Serum or Plasma', | Nucleated erythrocytes [#/volume] in Blood by Automated count', |
| Glucose [Mass/volume] in Serum or Plasma --2 hours post 75 g glucose PO', | Leukocytes [#/area] in Urine sediment by Microscopy high power field', | Hemoglobin A1/Hemoglobin.total in Blood', | Hepatitis C virus Ab [Presence] in Serum or Plasma by Immunoassay', | HIV 2 Ab [Presence] in Serum or Plasma by Immunoassay', | Nucleated erythrocytes [#/volume] in Blood', |
| Glucose [Mass/volume] in Serum or Plasma --2 hours post dose glucose', | Leukocytes [#/volume] corrected for nucleated erythrocytes in Blood by Automated count', | Hemoglobin A2.prime/Hemoglobin.total in Blood', | Hepatitis C virus Ab [Presence] in Serum', | HIV 2 Ab [Presence] in Serum'] | Nucleated erythrocytes/100 leukocytes [Ratio] in Blood by Automated count', |
| Glucose [Mass/volume] in Serum or Plasma --2 hours post XXX challenge', | Leukocytes [#/volume] in Blood by Automated count', | Hemoglobin A2/Hemoglobin.total in Blood by Chromatography column', | Hepatitis C virus Ab [Units/volume] in Serum by Immunoassay', | | Nucleated erythrocytes/100 leukocytes [Ratio] in Blood', |
| Glucose [Mass/volume] in Serum or Plasma --3 hours post 100 g glucose PO', | Leukocytes [#/volume] in Blood', | Hemoglobin A2/Hemoglobin.total in Blood by Electrophoresis', | Hepatitis C virus Ab Signal/Cutoff in Serum or Plasma by Immunoassay', | | Reticulocytes/100 erythrocytes in Blood by Automated count', |
| Glucose [Mass/volume] in Serum or Plasma --3 hours post dose glucose', | Leukocytes [#/volume] in Urine by Automated test strip', | Hemoglobin A2/Hemoglobin.total in Blood by HPLC', | Hepatitis C virus RNA [Units/volume] (viral load) in Serum or Plasma by NAA with probe detection', | | Reticulocytes/100 erythrocytes in Blood', |

| | | | |
|---|---|---|---|
| Glucose [Mass/volume] in Serum or Plasma --3 hours post XXX challenge', | Leukocytes [#/volume] in Urine by Test strip', | Hemoglobin A2/Hemoglobin.total in Blood', | Viral hepatitis C', |
| Glucose [Mass/volume] in Serum or Plasma --baseline', | Leukocytes [#/volume] in Urine', | Hemoglobin Barts/Hemoglobin.total in Blood', | |
| Glucose [Mass/volume] in Serum or Plasma --post XXX challenge', | Leukocytes [Presence] in Urine sediment by Light microscopy', | Hemoglobin C/Hemoglobin.total in Blood by Electrophoresis', | |
| Glucose [Mass/volume] in Serum or Plasma', | Leukocytes [Presence] in Urine', | Hemoglobin C/Hemoglobin.total in Blood', | |
| Glucose [Mass/volume] in Serum, Plasma or Blood', | Lymphocytes/100 leukocytes in Blood by Automated count', | Hemoglobin D/Hemoglobin.total in Blood', | |
| Glucose [Mass/volume] in Urine by Automated test strip', | Lymphocytes/100 leukocytes in Blood by Manual count', | Hemoglobin E/Hemoglobin.total in Blood', | |
| Glucose [Mass/volume] in Urine by Test strip', | Lymphocytes/100 leukocytes in Blood', | Hemoglobin F/Hemoglobin.total in Blood by Electrophoresis', | |
| Glucose [Mass/volume] in Urine', | Metamyelocytes/100 leukocytes in Blood by Manual count', | Hemoglobin F/Hemoglobin.total in Blood by Kleihauer-Betke method', | |
| Glucose [Moles/volume] in Capillary blood by Glucometer', | Monocytes/100 leukocytes in Blood by Automated count', | Hemoglobin F/Hemoglobin.total in Blood', | |
| Glucose [Moles/volume] in Serum or Plasma --1 hour post XXX challenge', | Monocytes/100 leukocytes in Blood by Manual count', | Hemoglobin G/Hemoglobin.total in Blood', | |
| Glucose [Presence] in Blood by Test strip', | Monocytes/100 leukocytes in Blood', | Hemoglobin Lepore/Hemoglobin.total in Blood', | |
| Glucose [Presence] in Urine by Automated test strip', | Myelocytes/100 leukocytes in Blood by Manual count', | Hemoglobin O - Arab/Hemoglobin.total in Blood', | |
| Glucose [Presence] in Urine by Test strip', | Neutrophils/100 leukocytes in Blood by Automated count', | Hemoglobin pattern [Interpretation] in Blood by Electrophoresis', | |
| Glucose [Presence] in Urine', | Neutrophils/100 leukocytes in Blood by Manual count', | Hemoglobin pattern [Interpretation] in Blood by HPLC Narrative', | |
| Glucose mean value [Mass/volume] in Blood Estimated from glycated hemoglobin', | Neutrophils/100 leukocytes in Blood', | Hemoglobin pattern [Interpretation] in Blood', | |
| Glucose.PO [Mass] of Dose', | Nucleated erythrocytes/100 leukocytes [Ratio] in Blood by Automated count', | Hemoglobin S [Presence] in Blood by Solubility test', | |
| Impaired glucose tolerance test', | Nucleated erythrocytes/100 leukocytes [Ratio] in Blood', | Hemoglobin S [Presence] in Blood', | |
| Impaired glucose tolerance'] | Polymorphonuclear cells/100 leukocytes in Blood', | Hemoglobin S/Hemoglobin.total in Blood by Electrophoresis', | |
| | Promyelocytes/100 leukocytes in Blood by Manual count', | Hemoglobin S/Hemoglobin.total in Blood by HPLC', | |
| | Segmented neutrophils/100 leukocytes in Blood by Automated count', | Hemoglobin S/Hemoglobin.total in Blood', | |

| | | |
|---|---|---|
| | Segmented neutrophils/100 leukocytes in Blood by Manual count', | Hemoglobin XXX/Hemoglobin.total in Blood by Electrophoresis', |
| | Segmented neutrophils/100 leukocytes in Blood', | Hemoglobin.other/Hemoglobin.total in Blood', |
| | Variant lymphocytes/100 leukocytes in Blood by Manual count', | Hemoglobinopathy'] |

#### Appendix E: Episode Time Distribution

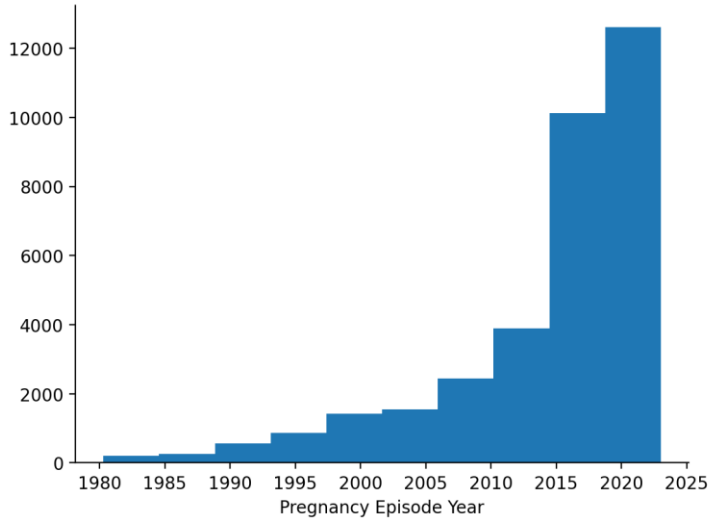

**Supplementary Figure 2** Histogram of time distribution (Pregnancy end time) for pregnancy episodes extracted. Only episodes within the recent 10 years were used (2015-2025)

#### Appendix F: Notable Features Table

After meeting with physicians to discuss the requirements and expectations from the clinical ratings, we additionally provide instructions provided to physicians for rating.

General Instructions:

Thank you very much for reviewing the features and providing feedback! For the six sheets in this document (corresponding to the six adverse outcomes), please review the Features and Values in columns B–G, and provide your comments in columns H–M. As a reminder, positive groups refer to participants who experienced the adverse outcome, while negative groups refer to those who did not. We greatly appreciate your time and insights. Your feedback will be essential in addressing reviewer comments as we prepare our revision for Nature Medicine.

Instructions for individual columns:

- **Column H: Expected/Unexpectedness of features.** Do you expect there are going to be differences in this value between positive and negative groups?
- **Column I Expected/Unexpected of numbers:** Do the numbers we are showing here make sense?

- **Column K: Used in Clinical Practice.** Imagine this value is abnormal, would this be a risk factor for the adverse outcome? Would this lead you to be concerned?
- **Column M: Used in Diagnosis?** Please check the checkbox it if this feature directly leads to diagnosis and should not be in a predictive model at all (e.g. Maternal Hypertension -> Preeclampsia)

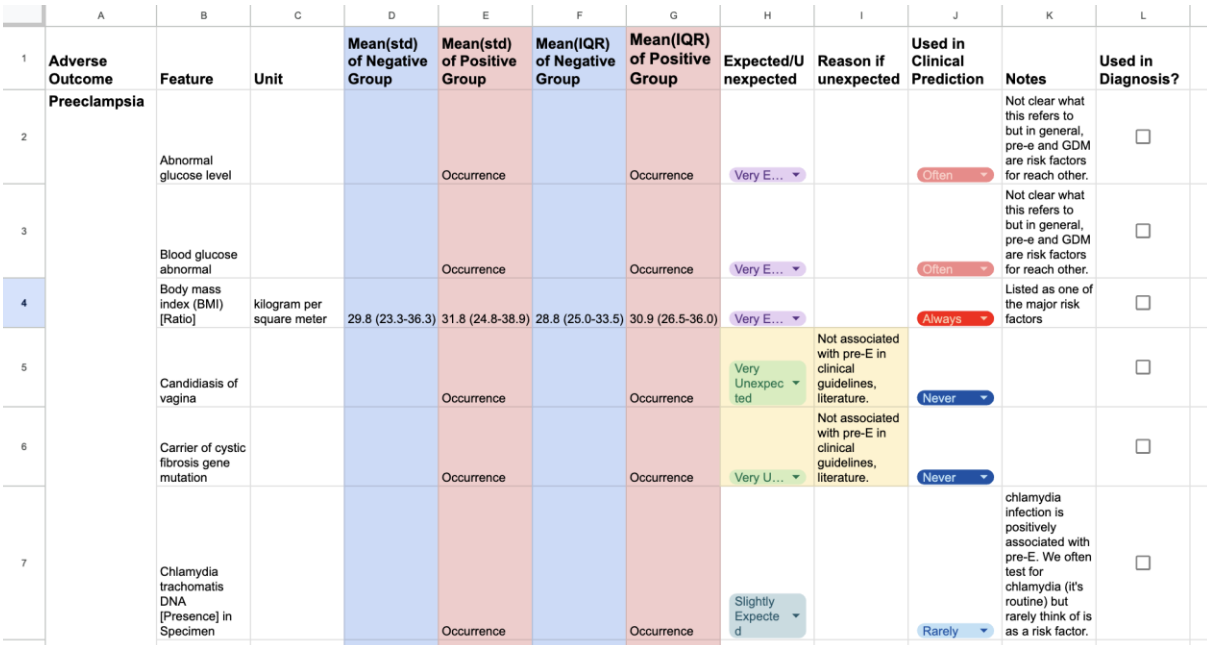

| | A | B | C | D | E | F | G | H | I | J | K | L |
|---|---|---|---|---|---|---|---|---|---|---|---|---|
| 1 | Adverse Outcome | Feature | Unit | Mean(std) of Negative Group | Mean(std) of Positive Group | Mean(IQR) of Negative Group | Mean(IQR) of Positive Group | Expected/Unexpected | Reason if unexpected | Used in Clinical Prediction | Notes | Used in Diagnosis? |
| 2 | Preeclampsia | Abnormal glucose level | | | Occurrence | | Occurrence | Very E… | | Often | Not clear what this refers to but in general, pre-e and GDM are risk factors for reach other. | ☐ |
| 3 | | Blood glucose abnormal | | | Occurrence | | Occurrence | Very E… | | Often | Not clear what this refers to but in general, pre-e and GDM are risk factors for reach other. | ☐ |
| 4 | | Body mass index (BMI) [Ratio] | kilogram per square meter | 29.8 (23.3-36.3) | 31.8 (24.8-38.9) | 28.8 (25.0-33.5) | 30.9 (26.5-36.0) | Very E… | | Always | Listed as one of the major risk factors | ☐ |
| 5 | | Candidiasis of vagina | | | Occurrence | | Occurrence | Very Unexpected | Not associated with pre-E in clinical guidelines, literature. | Never | | ☐ |
| 6 | | Carrier of cystic fibrosis gene mutation | | | Occurrence | | Occurrence | Very U… | Not associated with pre-E in clinical guidelines, literature. | Never | | ☐ |
| 7 | | Chlamydia trachomatis DNA [Presence] in Specimen | | | Occurrence | | Occurrence | Slightly Expected | | Rarely | chlamydia infection is positively associated with pre-E. We often test for chlamydia (it's routine) but rarely think of is as a risk factor. | ☐ |

**Supplementary Figure 3** Example table head from one of the sheets physician used to rate the features

Table: Full set of notable features

| adv | feature | unit | mean_std_negative | mean_std_positive | median_iqr_negative | median_iqr_positive | corr | xgb | lstm |
|---|---|---|---|---|---|---|---|---|---|
| preterm_labor | Heart rate | per minute | 85.3 (74.7-96.0) | 86.5 (75.9-97.1) | 85.2 (78.2-92.3) | 86.2 (79.5-93.4) | TRUE | TRUE | TRUE |
| preterm_labor | Glucose [Mass/volume] in Serum or Plasma | milligram per deciliter | 90.5 (76.4-104.6) | 89.8 (74.7-105.0) | 89.0 (81.5-97.5) | 88.0 (79.7-98.0) | FALSE | TRUE | FALSE |
| preterm_labor | Systolic blood pressure | millimeter mercury column | 113.1 (103.2-123.0) | 114.1 (103.3-124.8) | 112.3 (106.7-118.7) | 112.8 (106.8-119.0) | FALSE | TRUE | TRUE |
| preterm_labor | Lymphocytes/100 leukocytes in Blood | percent | 20.6 (12.7-28.5) | 20.9 (14.1-27.8) | 20.5 (16.0-25.3) | 20.6 (16.3-25.0) | FALSE | TRUE | FALSE |
| preterm_labor | Leukocytes [#/volume] in Urine | No matching concept | 3.6 (0.0-17.7) | 3.2 (0.0-13.2) | 0.0 (0.0-0.0) | 0.0 (0.0-0.0) | FALSE | TRUE | FALSE |
| preterm_labor | Glucose [Mass/volume] in Urine | milligram per deciliter | 8.9 (0.0-64.6) | 19.4 (0.0-113.3) | 0.0 (0.0-0.0) | 0.0 (0.0-0.0) | FALSE | FALSE | TRUE |
| preterm_labor | Diastolic blood pressure | millimeter mercury column | 69.9 (62.4-77.4) | 70.4 (62.5-78.4) | 69.6 (65.0-74.2) | 70.5 (65.3-75.0) | FALSE | FALSE | TRUE |
| preterm_labor | Hemoglobin [Mass/volume] in Blood | gram per liter | 119.4 (107.6-131.3) | 119.7 (107.7-131.7) | 120.0 (113.0-127.0) | 120.7 (112.8-127.5) | FALSE | FALSE | TRUE |
| preterm_labor | Protein [Mass/volume] in Urine | milligram per deciliter | 10.0 (0.0-45.2) | 18.2 (0.0-92.0) | 0.0 (0.0-10.0) | 0.0 (0.0-12.1) | FALSE | FALSE | TRUE |
| preterm_labor | Body mass index (BMI) [Ratio] | kilogram per square meter | 29.8 (23.3-36.3) | 30.0 (22.8-37.1) | 28.8 (25.0-33.5) | 28.8 (24.9-34.3) | FALSE | FALSE | TRUE |
| preterm_labor | Specific gravity of Urine | No matching concept | 1.0 (1.0-1.0) | 1.0 (1.0-1.0) | 1.0 (1.0-1.0) | 1.0 (1.0-1.0) | FALSE | FALSE | TRUE |

| | | | | | | | | | |
|---|---|---|---|---|---|---|---|---|---|
| depression_anxiety | Carbon dioxide [Partial pressure] in Arterial blood | millimeter mercury column | 33.5 (27.1-40.0) | 29.2 (20.8-37.6) | 32.0 (29.0-36.4) | 26.1 (23.7-31.2) | TRUE | FALSE | FALSE |
| depression_anxiety | Glucose [Mass/volume] in Serum or Plasma --2 hours post 100 g glucose PO | milligram per deciliter | 128.2 (102.8-153.6) | 147.3 (116.3-178.4) | 128.0 (111.0-144.0) | 146.0 (136.8-168.2) | TRUE | FALSE | FALSE |
| depression_anxiety | MCV [Entitic volume] | femtoliter | 88.2 (82.9-93.5) | 87.5 (82.2-92.9) | 88.6 (85.2-91.8) | 88.0 (84.4-91.0) | TRUE | FALSE | FALSE |
| depression_anxiety | Hemoglobin [Mass/volume] in Blood | gram per liter | 119.4 (107.6-131.3) | 119.9 (107.6-132.2) | 120.0 (113.0-127.0) | 120.7 (113.5-128.0) | FALSE | TRUE | FALSE |
| depression_anxiety | Glucose [Mass/volume] in Serum or Plasma --1 hour post dose glucose | milligram per deciliter | 116.1 (88.1-144.0) | 123.0 (89.2-156.9) | 113.0 (96.0-132.0) | 116.0 (96.0-142.0) | FALSE | TRUE | FALSE |
| depression_anxiety | Glucose [Mass/volume] in Serum or Plasma --1 hour post XXX challenge | milligram per deciliter | 113.2 (86.1-140.3) | 115.9 (84.5-147.3) | 110.0 (94.0-129.0) | 110.0 (91.0-138.5) | FALSE | TRUE | FALSE |
| depression_anxiety | Platelet mean volume [Entitic volume] in Blood | femtoliter | 10.0 (8.7-11.3) | 9.9 (8.6-11.2) | 10.1 (9.1-10.9) | 10.0 (9.1-10.8) | FALSE | TRUE | FALSE |
| depression_anxiety | Glucose [Mass/volume] in Serum or Plasma --1 hour post 50 g glucose PO | milligram per deciliter | 112.7 (86.5-138.8) | 114.2 (84.3-144.2) | 110.2 (94.0-129.9) | 113.5 (89.8-133.0) | FALSE | TRUE | FALSE |
| depression_anxiety | Choriogonadotropin [Units/volume] in Serum or Plasma | No matching concept | 38782.8 (3386.2-74179.4) | 39492.4 (6198.8-72786.0) | 29281.5 (8655.0-60748.8) | 32949.0 (12785.5-56104.3) | FALSE | FALSE | TRUE |
| depression_anxiety | Sodium [Moles/volume] in Serum or Plasma | millimole per liter | 136.9 (134.7-139.0) | 136.7 (134.6-138.7) | 137.0 (135.5-138.0) | 137.0 (135.1-138.0) | FALSE | FALSE | TRUE |
| depression_anxiety | Calcium [Mass/volume] in Serum or Plasma | milligram per deciliter | 9.1 (8.6-9.5) | 9.1 (8.6-9.5) | 9.1 (8.8-9.4) | 9.1 (8.8-9.4) | FALSE | FALSE | TRUE |
| depression_anxiety | Potassium [Moles/volume] in Serum or Plasma | millimole per liter | 3.9 (3.5-4.2) | 3.8 (3.5-4.2) | 3.8 (3.6-4.1) | 3.8 (3.6-4.0) | FALSE | FALSE | TRUE |
| depression_anxiety | Leukocytes [#/volume] in Urine | No matching concept | 3.6 (0.0-17.7) | 3.0 (0.0-12.1) | 0.0 (0.0-0.0) | 0.0 (0.0-0.0) | FALSE | FALSE | TRUE |
| depression_anxiety | Protein [Mass/volume] in Serum or Plasma | gram per deciliter | 6.9 (6.3-7.5) | 6.9 (6.3-7.5) | 6.9 (6.5-7.3) | 6.9 (6.5-7.2) | FALSE | FALSE | TRUE |
| depression_anxiety | Anion gap 3 in Serum or Plasma | No matching concept | 11.1 (8.6-13.6) | 11.2 (8.5-13.9) | 11.0 (9.0-13.0) | 11.0 (9.0-13.0) | FALSE | FALSE | TRUE |
| gestational_diabetes | Body mass index (BMI) [Ratio] | kilogram per square meter | 29.8 (23.3-36.3) | 33.0 (26.0-39.9) | 28.8 (25.0-33.5) | 32.1 (27.8-37.6) | TRUE | TRUE | TRUE |
| gestational_diabetes | Carbon dioxide [Partial pressure] in Blood | millimeter mercury column | 41.4 (33.8-49.0) | 33.7 (28.5-38.9) | 40.0 (37.0-46.5) | 35.0 (30.0-36.0) | TRUE | FALSE | FALSE |
| gestational_diabetes | Diastolic blood pressure | millimeter mercury column | 69.9 (62.4-77.4) | 72.1 (64.2-79.9) | 69.6 (65.0-74.2) | 72.0 (66.7-77.3) | TRUE | FALSE | TRUE |
| gestational_diabetes | Erythrocytes [#/volume] in Blood | million per microliter | 4.1 (3.7-4.5) | 4.2 (3.9-4.6) | 4.1 (3.8-4.3) | 4.2 (4.0-4.5) | TRUE | TRUE | TRUE |
| gestational_diabetes | Fasting glucose [Mass/volume] in Serum or Plasma | milligram per deciliter | 81.5 (73.5-89.4) | 88.4 (78.3-98.4) | 81.0 (76.0-87.0) | 87.0 (81.0-94.0) | TRUE | FALSE | FALSE |
| gestational_diabetes | Glucose [Mass/volume] in Blood | milligram per deciliter | 103.0 (75.4-130.6) | 120.5 (85.6-155.4) | 94.4 (85.0-112.0) | 118.0 (93.0-142.2) | TRUE | FALSE | FALSE |
| gestational_diabetes | Glucose [Mass/volume] in Serum or Plasma | milligram per deciliter | 90.5 (76.4-104.6) | 96.6 (80.1-113.1) | 89.0 (81.5-97.5) | 95.0 (86.0-106.0) | TRUE | TRUE | FALSE |
| gestational_diabetes | Glucose [Mass/volume] in Serum or Plasma --1 hour post 100 g glucose PO | milligram per deciliter | 135.6 (102.6-168.7) | 172.0 (147.0-197.0) | 138.0 (109.0-160.0) | 171.5 (148.7-190.2) | TRUE | FALSE | FALSE |
| gestational_diabetes | Glucose [Mass/volume] in Serum or Plasma --1 hour post 50 g glucose PO | milligram per deciliter | 112.7 (86.5-138.8) | 148.6 (122.2-175.0) | 110.2 (94.0-129.9) | 147.0 (134.8-168.8) | TRUE | TRUE | FALSE |
| gestational_diabetes | Glucose [Mass/volume] in Serum or Plasma --1 hour post XXX challenge | milligram per deciliter | 113.2 (86.1-140.3) | 147.9 (116.1-179.6) | 110.0 (94.0-129.0) | 150.0 (127.0-169.9) | TRUE | FALSE | FALSE |
| gestational_diabetes | Glucose [Mass/volume] in Serum or Plasma --1 hour post dose glucose | milligram per deciliter | 116.1 (88.1-144.0) | 157.4 (123.2-191.6) | 113.0 (96.0-132.0) | 159.0 (135.5-183.0) | TRUE | TRUE | FALSE |
| gestational_diabetes | Glucose [Mass/volume] in Serum or Plasma --2 hours post 100 g glucose PO | milligram per deciliter | 128.2 (102.8-153.6) | 161.5 (129.2-193.7) | 128.0 (111.0-144.0) | 160.0 (134.0-190.0) | TRUE | FALSE | FALSE |

| | | | | | | | | | |
|---|---|---|---|---|---|---|---|---|---|
| gestational_diabetes | Glucose [Mass/volume] in Serum or Plasma --2 hours post dose glucose | milligram per deciliter | 126.0 (100.1-151.8) | 154.4 (125.6-183.2) | 125.5 (107.0-144.0) | 154.2 (136.4-174.8) | TRUE | FALSE | FALSE |
| gestational_diabetes | Glucose [Mass/volume] in Urine | milligram per deciliter | 8.9 (0.0-64.6) | 30.8 (0.0-135.3) | 0.0 (0.0-0.0) | 0.0 (0.0-0.0) | TRUE | FALSE | FALSE |
| gestational_diabetes | Glucose mean value [Mass/volume] in Blood Estimated from glycated hemoglobin | milligram per deciliter | 105.2 (91.3-119.2) | 111.0 (98.4-123.6) | 103.0 (97.0-111.0) | 111.0 (100.0-120.0) | TRUE | FALSE | FALSE |
| gestational_diabetes | Heart rate | per minute | 85.3 (74.7-96.0) | 87.0 (76.6-97.4) | 85.2 (78.2-92.3) | 86.6 (80.0-93.3) | TRUE | FALSE | FALSE |
| gestational_diabetes | Hematocrit [Volume Fraction] of Blood | percent | 35.8 (32.6-39.1) | 36.8 (33.7-39.9) | 36.0 (33.9-37.9) | 36.8 (35.0-38.7) | TRUE | TRUE | FALSE |
| gestational_diabetes | Hemoglobin [Mass/volume] in Blood | gram per liter | 119.4 (107.6-131.3) | 122.9 (111.2-134.6) | 120.0 (113.0-127.0) | 123.0 (115.3-131.0) | TRUE | TRUE | TRUE |
| gestational_diabetes | Leukocytes [#/volume] in Blood | thousand per microliter | 9.1 (6.8-11.3) | 9.4 (7.1-11.8) | 8.9 (7.5-10.5) | 9.4 (7.8-10.9) | TRUE | FALSE | TRUE |
| gestational_diabetes | Lymphocytes [#/volume] in Blood | thousand per microliter | 1.9 (1.3-2.5) | 2.1 (1.4-2.8) | 1.8 (1.5-2.2) | 2.0 (1.6-2.5) | TRUE | FALSE | FALSE |
| gestational_diabetes | MCH [Entitic mass] | picogram | 29.5 (27.4-31.7) | 29.2 (27.1-31.4) | 29.8 (28.4-31.0) | 29.5 (28.0-30.7) | TRUE | FALSE | TRUE |
| gestational_diabetes | MCV [Entitic volume] | femtoliter | 88.2 (82.9-93.5) | 87.2 (81.9-92.6) | 88.6 (85.2-91.8) | 87.7 (84.0-91.0) | TRUE | FALSE | FALSE |
| gestational_diabetes | Monocytes/100 leukocytes in Blood | percent | 6.5 (4.8-8.1) | 6.2 (4.6-7.8) | 6.3 (5.3-7.5) | 6.0 (5.0-7.0) | TRUE | FALSE | FALSE |
| gestational_diabetes | Platelets [#/volume] in Blood | thousand per microliter | 248.2 (192.2-304.3) | 263.4 (201.7-325.0) | 243.7 (209.0-283.0) | 258.4 (221.0-302.1) | TRUE | FALSE | TRUE |
| gestational_diabetes | Systolic blood pressure | millimeter mercury column | 113.1 (103.2-123.0) | 116.7 (105.9-127.5) | 112.3 (106.7-118.7) | 116.1 (109.4-123.3) | TRUE | FALSE | TRUE |
| gestational_diabetes | Erythrocyte distribution width [Ratio] | percent | 13.5 (12.5-14.5) | 13.6 (12.5-14.6) | 13.3 (12.8-14.0) | 13.4 (12.9-14.2) | FALSE | FALSE | TRUE |
| pre_eclampsia | Bilirubin.indirect [Mass/volume] in Serum or Plasma | milligram per deciliter | 0.3 (0.2-0.5) | 0.2 (0.1-0.4) | 0.3 (0.2-0.4) | 0.2 (0.1-0.3) | TRUE | FALSE | FALSE |
| pre_eclampsia | Body mass index (BMI) [Ratio] | kilogram per square meter | 29.8 (23.3-36.3) | 31.8 (24.8-38.9) | 28.8 (25.0-33.5) | 30.9 (26.5-36.0) | TRUE | FALSE | TRUE |
| pre_eclampsia | Diastolic blood pressure | millimeter mercury column | 69.9 (62.4-77.4) | 73.5 (66.3-80.8) | 69.6 (65.0-74.2) | 73.5 (68.7-78.4) | TRUE | TRUE | TRUE |
| pre_eclampsia | Erythrocyte distribution width [Ratio] | percent | 13.5 (12.5-14.5) | 13.7 (12.7-14.8) | 13.3 (12.8-14.0) | 13.5 (12.9-14.3) | TRUE | FALSE | FALSE |
| pre_eclampsia | Fasting glucose [Mass/volume] in Serum or Plasma | milligram per deciliter | 81.5 (73.5-89.4) | 84.9 (75.4-94.5) | 81.0 (76.0-87.0) | 84.0 (78.0-91.9) | TRUE | FALSE | FALSE |
| pre_eclampsia | Glucose [Mass/volume] in Serum or Plasma --1 hour post 50 g glucose PO | milligram per deciliter | 112.7 (86.5-138.8) | 123.9 (94.9-153.0) | 110.2 (94.0-129.9) | 122.0 (100.0-141.2) | TRUE | FALSE | FALSE |
| pre_eclampsia | Glucose [Mass/volume] in Serum or Plasma --1 hour post dose glucose | milligram per deciliter | 116.1 (88.1-144.0) | 131.8 (95.4-168.3) | 113.0 (96.0-132.0) | 126.5 (103.0-155.9) | TRUE | FALSE | FALSE |
| pre_eclampsia | Glucose [Mass/volume] in Serum or Plasma --2 hours post 100 g glucose PO | milligram per deciliter | 128.2 (102.8-153.6) | 152.2 (118.0-186.4) | 128.0 (111.0-144.0) | 141.0 (127.0-179.0) | TRUE | FALSE | FALSE |
| pre_eclampsia | Glucose [Mass/volume] in Urine | milligram per deciliter | 8.9 (0.0-64.6) | 35.5 (0.0-158.3) | 0.0 (0.0-0.0) | 0.0 (0.0-0.0) | TRUE | FALSE | FALSE |
| pre_eclampsia | Glucose mean value [Mass/volume] in Blood Estimated from glycated hemoglobin | milligram per deciliter | 105.2 (91.3-119.2) | 113.4 (97.4-129.4) | 103.0 (97.0-111.0) | 111.0 (103.0-120.7) | TRUE | FALSE | FALSE |
| pre_eclampsia | Heart rate | per minute | 85.3 (74.7-96.0) | 87.5 (76.7-98.3) | 85.2 (78.2-92.3) | 87.2 (80.0-94.7) | TRUE | FALSE | TRUE |
| pre_eclampsia | Leukocytes [#/volume] in Blood | thousand per microliter | 9.1 (6.8-11.3) | 9.5 (7.2-11.7) | 8.9 (7.5-10.5) | 9.4 (8.0-10.9) | TRUE | FALSE | FALSE |
| pre_eclampsia | Platelet mean volume [Entitic volume] in Blood | femtoliter | 10.0 (8.7-11.3) | 9.8 (8.4-11.3) | 10.1 (9.1-10.9) | 9.8 (8.8-10.9) | TRUE | FALSE | FALSE |
| pre_eclampsia | Sodium [Moles/volume] in Serum or Plasma | millimole per liter | 136.9 (134.7-139.0) | 136.3 (134.2-138.4) | 137.0 (135.5-138.0) | 136.3 (135.0-138.0) | TRUE | FALSE | FALSE |

| | | | | | | | | | |
|---|---|---|---|---|---|---|---|---|---|
| pre_eclampsia | Systolic blood pressure | millimeter mercury column | 113.1 (103.2-123.0) | 119.2 (109.4-129.0) | 112.3 (106.7-118.7) | 118.5 (112.3-125.4) | TRUE | TRUE | TRUE |
| pre_eclampsia | Thyroxine (T4) free [Mass/volume] in Serum or Plasma | nanogram per deciliter | 1.1 (0.8-1.3) | 1.0 (0.7-1.3) | 1.1 (0.9-1.2) | 1.0 (0.8-1.1) | TRUE | FALSE | FALSE |
| pre_eclampsia | Erythrocyte distribution width [Entitic volume] | femtoliter | 42.9 (39.7-46.2) | 42.2 (38.0-46.4) | 42.5 (40.7-44.9) | 41.7 (39.3-45.5) | FALSE | TRUE | FALSE |
| pre_eclampsia | Bacteria [#/area] in Urine sediment | | | Occurrence | | Occurrence | FALSE | TRUE | FALSE |
| pre_eclampsia | pH of Urine | No matching concept | 6.4 (5.8-7.1) | 6.4 (5.8-7.0) | 6.5 (6.0-6.9) | 6.3 (6.0-6.9) | FALSE | FALSE | TRUE |
| pre_eclampsia | Specific gravity of Urine | No matching concept | 1.0 (1.0-1.0) | 1.0 (1.0-1.0) | 1.0 (1.0-1.0) | 1.0 (1.0-1.0) | FALSE | FALSE | TRUE |
| miscarriage | Cholesterol non HDL [Mass/volume] in Serum or Plasma | milligram per deciliter | 118.0 (84.1-151.9) | 158.5 (123.2-193.8) | 114.0 (95.0-136.5) | 157.5 (132.5-183.5) | TRUE | FALSE | FALSE |
| miscarriage | Diastolic blood pressure | millimeter mercury column | 69.9 (62.4-77.4) | 73.2 (64.8-81.7) | 69.6 (65.0-74.2) | 72.6 (67.9-78.2) | TRUE | TRUE | FALSE |
| miscarriage | Erythrocytes [#/volume] in Blood | million per microliter | 4.1 (3.7-4.5) | 4.3 (3.8-4.8) | 4.1 (3.8-4.3) | 4.3 (4.0-4.6) | TRUE | TRUE | FALSE |
| miscarriage | Hematocrit [Volume Fraction] of Blood | percent | 35.8 (32.6-39.1) | 37.1 (33.3-41.0) | 36.0 (33.9-37.9) | 37.5 (34.6-39.3) | TRUE | TRUE | FALSE |
| miscarriage | Hemoglobin [Mass/volume] in Blood | gram per liter | 119.4 (107.6-131.3) | 123.2 (109.2-137.2) | 120.0 (113.0-127.0) | 123.5 (114.0-132.0) | TRUE | FALSE | FALSE |
| miscarriage | MCH [Entitic mass] | picogram | 29.5 (27.4-31.7) | 28.7 (26.6-30.8) | 29.8 (28.4-31.0) | 29.0 (27.5-30.1) | TRUE | FALSE | FALSE |
| miscarriage | MCV [Entitic volume] | femtoliter | 88.2 (82.9-93.5) | 86.0 (80.7-91.4) | 88.6 (85.2-91.8) | 86.3 (83.8-89.2) | TRUE | TRUE | FALSE |
| miscarriage | Systolic blood pressure | millimeter mercury column | 113.1 (103.2-123.0) | 116.8 (105.2-128.4) | 112.3 (106.7-118.7) | 115.9 (109.0-123.5) | TRUE | FALSE | FALSE |
| miscarriage | Glucose [Mass/volume] in Serum or Plasma --1 hour post 50 g glucose PO | milligram per deciliter | 112.7 (86.5-138.8) | 127.7 (106.4-148.9) | 110.2 (94.0-129.9) | 132.0 (118.8-138.5) | FALSE | TRUE | FALSE |
| miscarriage | Leukocytes [#/volume] in Blood | thousand per microliter | 9.1 (6.8-11.3) | 8.2 (5.7-10.6) | 8.9 (7.5-10.5) | 8.0 (6.4-9.5) | FALSE | TRUE | FALSE |
| miscarriage | Heart rate | per minute | 85.3 (74.7-96.0) | 84.7 (73.3-96.2) | 85.2 (78.2-92.3) | 84.0 (77.0-91.6) | FALSE | TRUE | TRUE |
| miscarriage | Body temperature | degree Celsius | 36.7 (36.5-37.0) | 36.8 (36.5-37.0) | 36.7 (36.6-36.9) | 36.8 (36.6-36.9) | FALSE | FALSE | TRUE |
| miscarriage | Specific gravity of Urine | No matching concept | 1.0 (1.0-1.0) | 1.0 (1.0-1.0) | 1.0 (1.0-1.0) | 1.0 (1.0-1.0) | FALSE | FALSE | TRUE |
| preterm_labor | Abdominal pain | | | Occurrence | | Occurrence | TRUE | TRUE | FALSE |
| preterm_labor | Cervical incompetence | | | Occurrence | | Occurrence | TRUE | FALSE | FALSE |
| preterm_labor | Ketones [Presence] in Urine | No matching concept | 77741.8 (0.0-944471.0) | 0.1 (0.0-1.3) | 0.0 (0.0-0.0) | 0.0 (0.0-0.0) | TRUE | FALSE | FALSE |
| preterm_labor | Urobilinogen [Presence] in Urine | No matching concept | 0.3 (0.0-1.1) | 0.5 (0.0-1.4) | 0.0 (0.0-0.0) | 0.0 (0.0-0.1) | TRUE | FALSE | FALSE |
| preterm_labor | Bilirubin.total [Presence] in Urine | No matching concept | 0.0 (0.0-0.0) | 0.0 (0.0-0.0) | 0.0 (0.0-0.0) | 0.0 (0.0-0.0) | FALSE | FALSE | TRUE |
| depression_anxiety | Anemia in mother complicating childbirth | | | Occurrence | | Occurrence | TRUE | FALSE | FALSE |
| depression_anxiety | Cord entanglement without compression | | | Occurrence | | Occurrence | TRUE | FALSE | FALSE |
| depression_anxiety | Group B streptococcus infection in mother complicating childbirth | | | Occurrence | | Occurrence | TRUE | FALSE | FALSE |
| depression_anxiety | Mild hyperemesis gravidarum | | | Occurrence | | Occurrence | FALSE | FALSE | TRUE |
| depression_anxiety | Urobilinogen [Presence] in Urine | No matching concept | 0.3 (0.0-1.1) | 0.4 (0.0-1.2) | 0.0 (0.0-0.0) | 0.0 (0.0-0.0) | FALSE | FALSE | TRUE |
| gestational_diabetes | Blood glucose abnormal | | | Occurrence | | Occurrence | TRUE | FALSE | FALSE |

| | | | | | | | | | |
|---|---|---|---|---|---|---|---|---|---|
| gestational_diabetes | Morbid obesity | | | Occurrence | | Occurrence | TRUE | FALSE | FALSE |
| gestational_diabetes | Polycystic ovary syndrome | | | Occurrence | | Occurrence | TRUE | FALSE | FALSE |
| gestational_diabetes | Worried well | | | Occurrence | | Occurrence | TRUE | FALSE | FALSE |
| pre_eclampsia | Abnormal glucose level | | | Occurrence | | Occurrence | TRUE | FALSE | FALSE |
| pre_eclampsia | Blood glucose abnormal | | | Occurrence | | Occurrence | TRUE | FALSE | FALSE |
| pre_eclampsia | Carrier of cystic fibrosis gene mutation | | | Occurrence | | Occurrence | TRUE | FALSE | FALSE |
| pre_eclampsia | Chlamydia trachomatis DNA [Presence] in Specimen | | | Occurrence | | Occurrence | TRUE | FALSE | FALSE |
| pre_eclampsia | Kidney disease | | | Occurrence | | Occurrence | TRUE | FALSE | FALSE |
| pre_eclampsia | Morbid obesity | | | Occurrence | | Occurrence | TRUE | FALSE | FALSE |
| pre_eclampsia | Obesity | | | Occurrence | | Occurrence | TRUE | FALSE | FALSE |
| pre_eclampsia | Proteinuria | | | Occurrence | | Occurrence | TRUE | FALSE | FALSE |
| pre_eclampsia | Suspected fetal abnormality affecting management of mother | | | Occurrence | | Occurrence | TRUE | FALSE | FALSE |
| pre_eclampsia | Systemic lupus erythematosus | | | Occurrence | | Occurrence | TRUE | FALSE | FALSE |
| pre_eclampsia | Hemoglobin [Presence] in Urine | No matching concept | 0.0 (0.0-1.3) | 0.0 (0.0-0.0) | 0.0 (0.0-0.0) | 0.0 (0.0-0.0) | FALSE | FALSE | TRUE |
| pre_eclampsia | Leukocyte esterase [Presence] in Urine | | | Occurrence | | Occurrence | FALSE | FALSE | TRUE |
| eclampsia | Exacerbation of intermittent asthma | | | Occurrence | | Occurrence | TRUE | FALSE | FALSE |
| eclampsia | Fibronectin.fetal [Presence] in Specimen | No matching concept | -0.8 (0.0--0.2) | 1.0 (1.0-1.0) | -1.0 (-1.0--1.0) | 1.0 (1.0-1.0) | TRUE | FALSE | FALSE |

### Appendix G: All Clinical Ratings

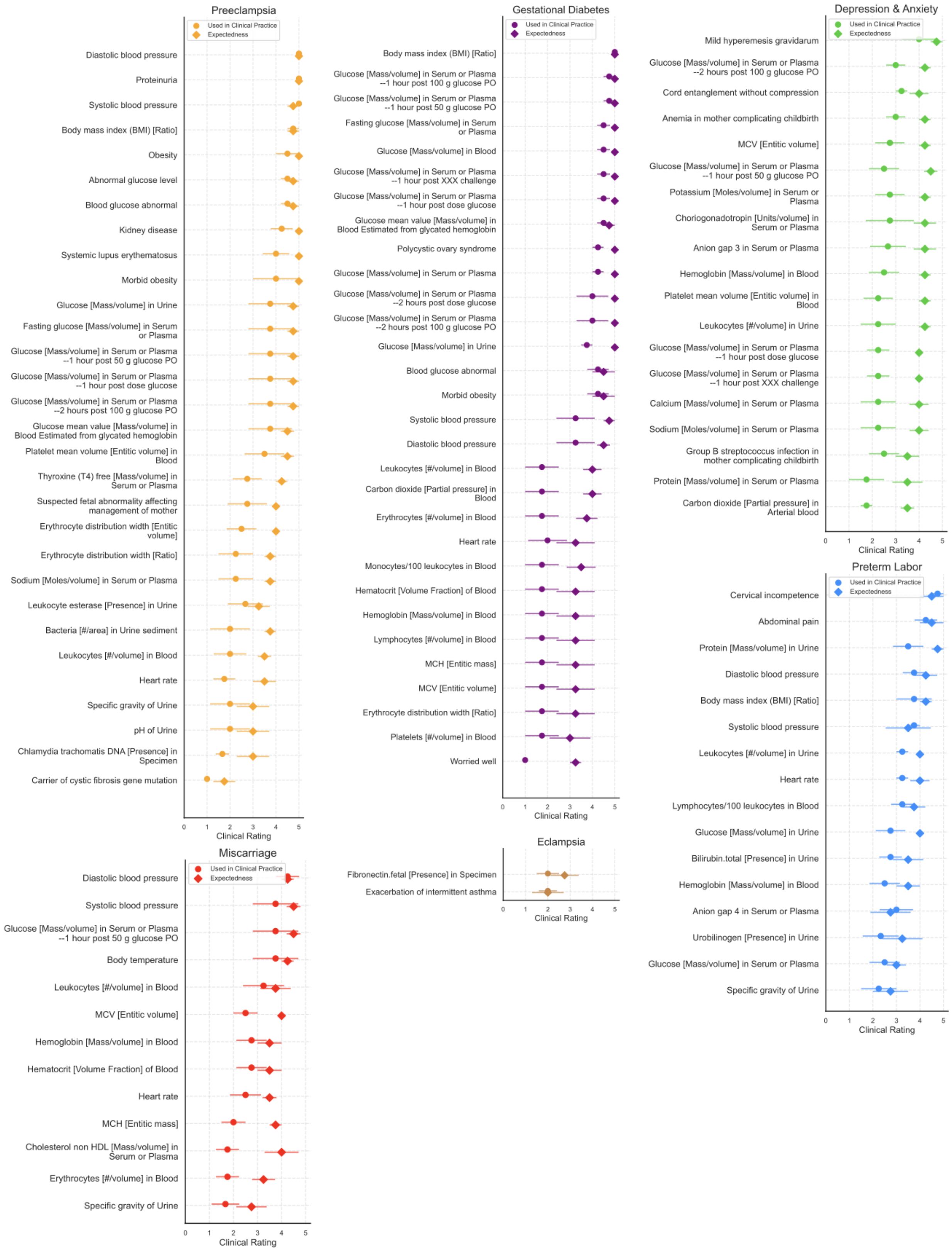

### Appendix H: All Feature Comparisons

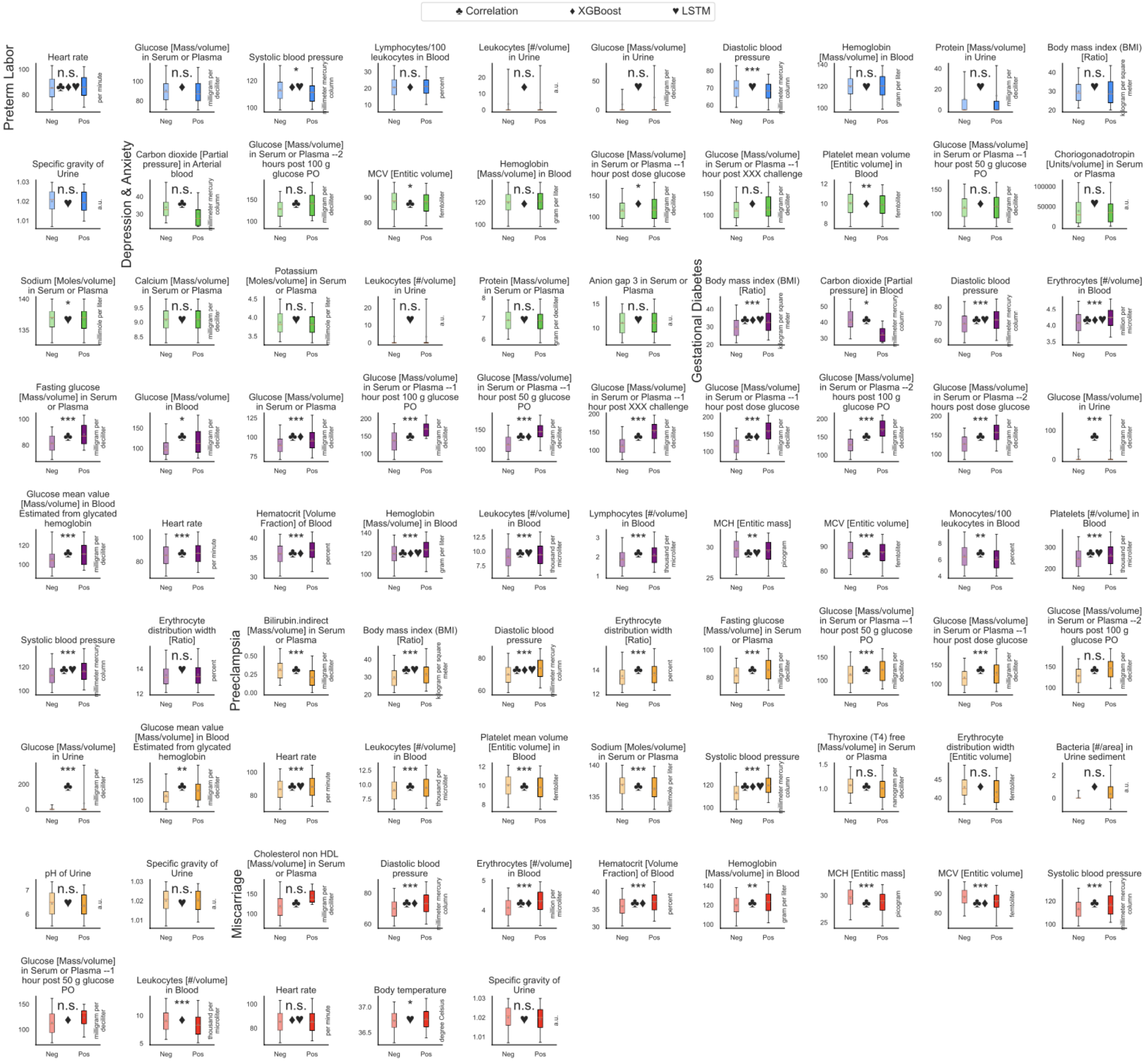

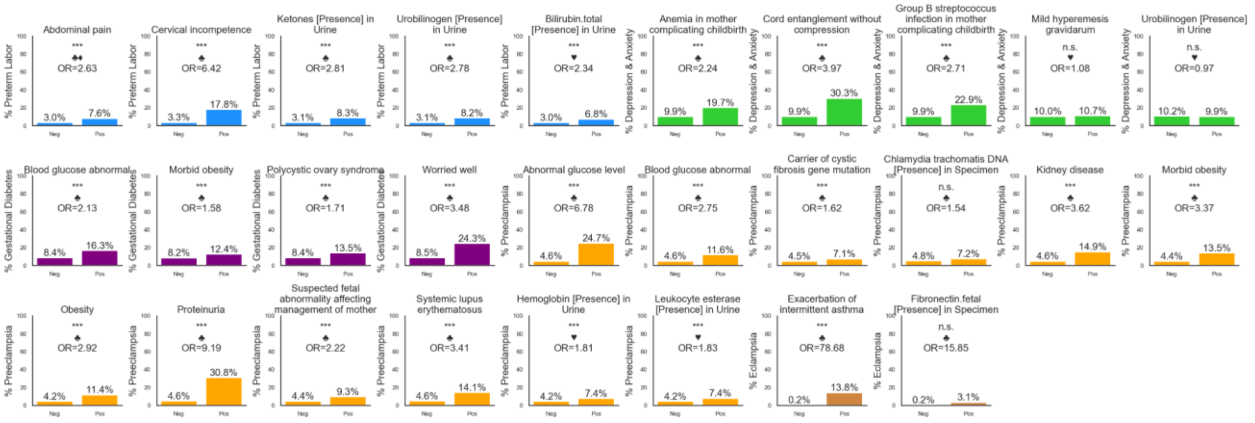

### Appendix I: Full Correlation Heatmap

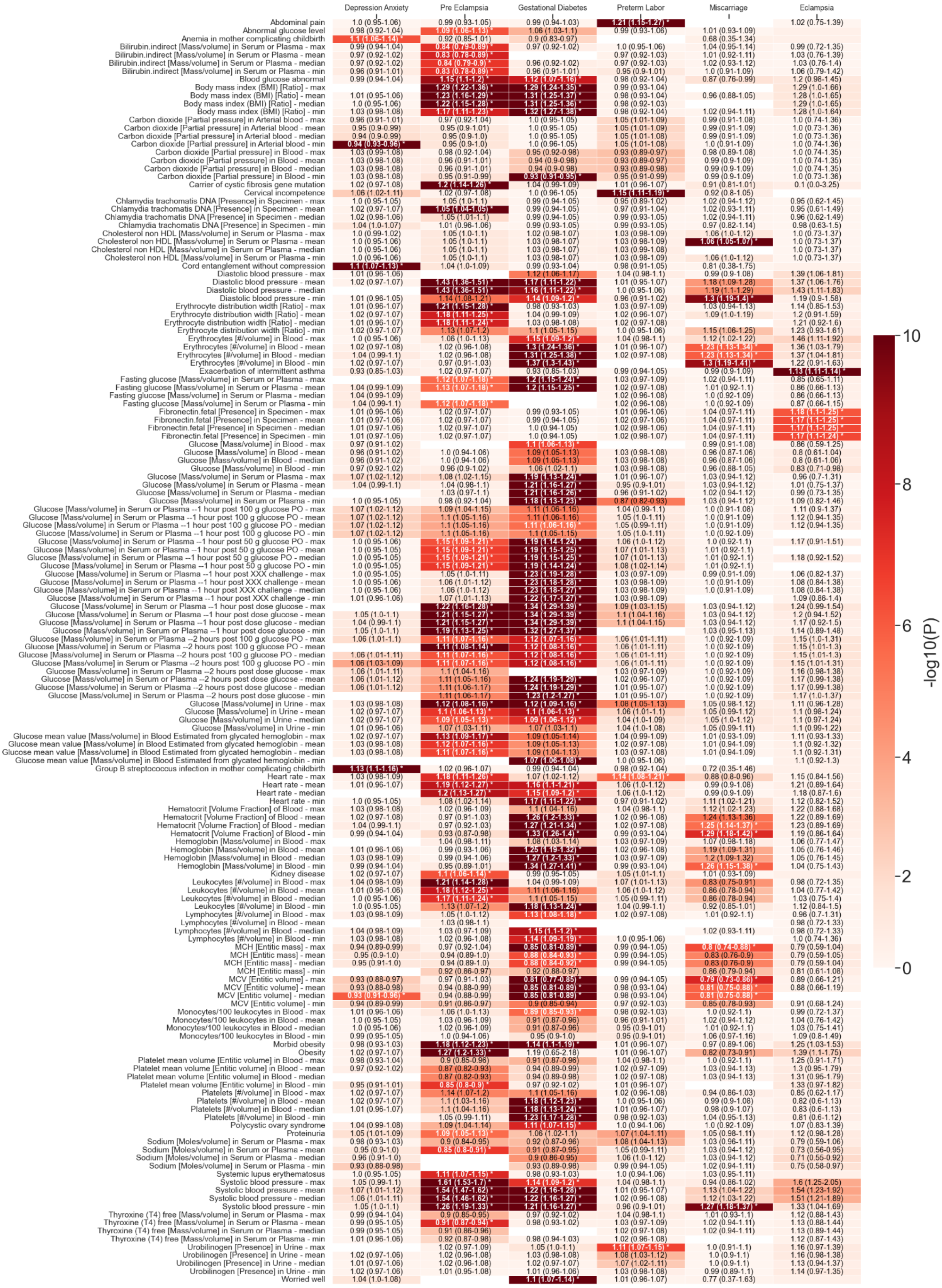

#### Appendix J: XGBoost Feature Importance is Associated with Odds Ratio

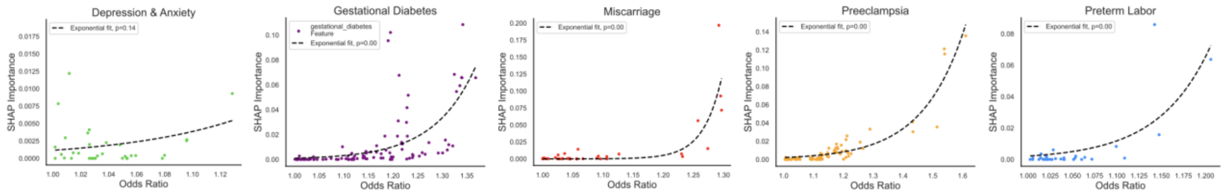

#### Appendix K: LSTM Feature Importance is Associated with Occurrence

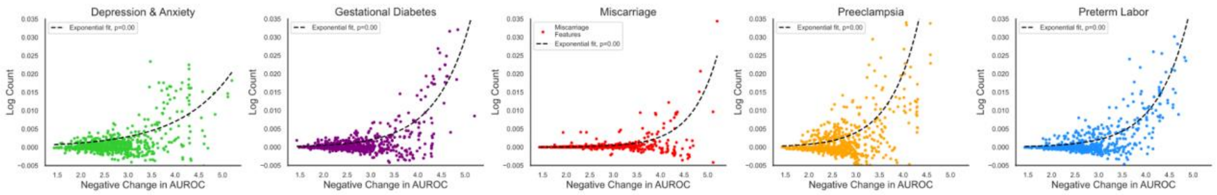
